# Individualised evoked response detection based on the spectral noise colour

**DOI:** 10.64898/2026.04.11.26350685

**Authors:** Jaime A. Undurraga, Michael Chesnaye, David Simpson, Søren Laugesen

## Abstract

Statistical inference in neurophysiological recordings is fundamentally challenged by the presence of coloured—1/*f* -like—background activity, which introduces temporal dependencies and violates the assumptions underlying many conventional detection methods. In both electroencephography (EEG) and magnetoencephography (MEG), this mismatch can lead to biased estimates of statistical significance and reduced robustness across recordings and individuals. Here, we present Fmpi, a general framework for statistical inference that explicitly accounts for the spectral colour and temporal dynamics of background noise by deriving analytical estimates of the effective degrees of freedom directly from the power spectral density of the data. We demonstrate the utility of the framework in the context of auditory evoked potential (AEP) detection using extensive simulations and large-scale human EEG datasets including brainstem, steady-state, and cortical responses recorded in adults and infants. Across both stationary and non-stationary noise conditions, the proposed method provides well-calibrated statistical inference, with sensitivity and specificity comparable or superior to state-of-the-art bootstrapped approaches, while operating at a fraction of the computational cost. By adapting detection thresholds to the noise characteristics of individual recordings, Fmpi reduces bias associated with population-based assumptions and improves detection efficiency. In addition, the framework enables prediction of the number of observations required for reliable detection, supporting a principled futility-stopping criterion that substantially reduces recording time without compromising diagnostic performance.

Although demonstrated here for AEP detection, the proposed approach is broadly applicable to other inference problems in neurophysiology where coloured noise and temporal correlations play a central role.

**Highlights:**

- Analytical inference under coloured (1/*f*) neural noise
- Individualised DOF from power spectral density
- Matches bootstrap performance at low computational cost
- Improved specificity with well-calibrated sensitivity
- Enables futility stopping to reduce recording time

## 1 Introduction

When we hear a sound, look at objects, move, or touch something, millions of neurons are activated, generating electromagnetic signals at different stages throughout the nervous system. These electromagnetic signals are known as evoked potentials (EPs) and play a crucial role in clinical diagnosis, intraoperative testing and neuroscience research. EPs can be detected non-invasively using extracorporeal techniques such as magneto- and electro-encephalography (MEG and EEG), invasively through sensors placed in direct contact with neural tissue (e.g., electrocorticography), or indirectly via stimulus-evoked hemodynamic responses measured with functional near-infrared spectroscopy (fNIRS).

Clinically, EPs are used to assess and diagnose sensory-neural dysfunction along the neural pathway for different sensory modalities. For example, auditory brainstem responses (ABRs)—an early auditory evoked potential (AEP) reflecting hearing function from the auditory nerve to the midbrain—play a critical role in newborn hearing screening (*World report on hearing* 2021), the diagnosis of auditory neuropathy (Starr et al., 1996) (a dysfunction in the transmission of action potentials along the auditory nerve), and intraoperative monitoring of auditory function (Matthies and Samii, 1997). Similarly, cortical AEPs—reflecting neural activity evoked by sounds at the cortex—play an important role for clinicians and primary caregivers in determining strategies to manage hearing loss (Mehta et al., 2017).

Beyond the auditory domain, visual evoked potentials (VEPs), somatosensory evoked potentials (SEPs), and vestibular evoked myogenic potentials (VEMPs) are used clinically for the diagnosis of multiple sclerosis, intraoperative monitoring, diagnosis of optic nerve tumors or optic neuropathy, and assessment of vestibular function, among other applications (Lascano et al., 2017).

Regardless of the sensory modality being tested, the measurement of an EP is, in principle, relatively straightforward. One or several types of stimuli are presented repeatedly—ranging from a few hundred repetitions (e.g., for cortical EPs) to several thousand (e.g., for brainstem EPs). The data are then filtered to reduce noise outside the EP bandwidth. Finally, data blocks aligned to each stimulus onset (epochs or trials) are averaged to further reduce noise within the bandwidth of interest.

The stimulus presentation rate—or, more generally, the rate of the specific cue within the stimulus that evokes the EP response—plays a crucial role in determining the type of neural response elicited. For example, low presentation rates—relative to the time course of the response—are usually employed when the morphology of the EP is of interest. Low presentation rates evoke a transient-like EP, where the relatively long interval between presentations prevents overlap of consecutive EPs—e.g., ABRs (Jewett and Williston, 1971), auditory change complex (ACC) responses (Martin and Boothroyd, 2000), VEPs (Cigánek, 1961; Taylor and McCulloch, 1992; Aminoff and Goodin, 1994), VEMPs (Colebatch and Halmagyi, 1992)—and minimise reduced neural responsiveness due to prior excitation (rate adaptation). When the stimulus cue evoking the EP is presented periodically and at higher rates, the morphology of the EP will be determined by the extent of overlap between consecutive responses, and it will follow—entrain to—such periodicity, generating the so-called steady-state EP, e.g., auditory steady-state responses (ASSRs) (Galambos et al., 1981; Picton et al., 2003), interaural phase modulation following responses (IPM-FRs) and interaural time difference following responses (ITD-FRs) (Undurraga et al., 2016; Undurraga et al., 2024), auditory change following responses (AC-FRs) (Undurraga et al., 2020a), steady-state VEPs (Regan, 1977; Norcia et al., 2015), somatosensory steady-state responses (Snyder, 1992), or steady-state VEMPs (Bell et al., 2010).

Despite these relatively straightforward approaches to eliciting conventional EPs, accurately and efficiently determining their presence or absence remains a challenging topic. This is because neurophysiological recordings such as EEG, MEG, and fNIRS are inherently dominated by background activity exhibiting strong temporal correlations and a characteristic 1/*f* -like spectral structure. This coloured noise violates the assumptions of independence and stationarity that underpin many commonly used statistical inference frameworks.

Decades of research have led to the development of multiple approaches to detect neurophysiological responses in the time domain, frequency domain, or both domains. These include statistical (Wastell, 1977; Mason et al., 1977; Wicke et al., 1978; Elberling and Don, 1984; Don et al., 1984; Arnold, 1985; Victor and Mast, 1991; Champlin, 1992; Don and Elberling, 1994; Henning and Husar, 1995; Valdes et al., 1997; Liavas et al., 1998; Stürzebecher et al., 1999; Cebulla et al., 2006; Silva, 2009; Golding et al., 2009; Stürzebecher and Cebulla, 2013; Cebulla and Stürzebecher, 2015; Patel et al., 2017; Chesnaye et al., 2018; Bardy et al., 2020; Derzsi, 2021; Chesnaye et al., 2021a; Chesnaye et al., 2023) and machine-learning approaches (Alpsan, 1991; Acır et al., 2006; Davey et al., 2007; Parvar et al., 2015; McKearney and MacKinnon, 2019; Zhu et al., 2019; Yperman et al., 2020; Xing et al., 2020; McKearney et al., 2022).

Whilst these approaches have been useful for research and clinical diagnosis, their full potential has not yet been realised, partly due to the assumptions and constraints inherent to each approach. For example, assumptions about the properties of the underlying background noise (BN) in the recordings are often violated in practice (Lv et al., 2007; Maris and Oostenveld, 2007; Chesnaye et al., 2021b). The need for feature selection (e.g., the specific number and type of features used by statistical or machine learning approaches) varies depending on the type of EPs, and sub-optimal choices may lead to a loss of information, prolonged detection times, and reduced correct response detection rates—specificity. Additionally, the requirement for high computational power (e.g., bootstrapping methods) can hinder both real-time and clinical implementation.

To ensure sufficient test sensitivity—here defined clinically as the correct identification of the absence of a neurophysiological response—certain detection criteria rely on the population-level distribution of a given test statistic (Elberling and Don, 1984; Don et al., 1984; Don and Elberling, 1994; Stürzebecher and Cebulla, 2013; Cebulla and Stürzebecher, 2015; Lightfoot et al., 2023) rather than the specific statistical characteristics of an individual recording. Other approaches estimate response detection thresholds from features extracted directly from the recording, albeit informed by fixed population-derived features (Golding et al., 2009; Chesnaye et al., 2018; Bardy et al., 2020; Chesnaye et al., 2021a; McKearney et al., 2022). Whilst these strategies typically ensure a minimum sensitivity of 95% in clinical contexts, the lack or limited degree of individualisation often results in elevated detection thresholds and unnecessarily prolonged test durations.

Improvements in the estimation of response detection thresholds for each individual recording can be expected to lead to more accurate and efficient diagnoses. This is particularly important in the context of hearing, as health systems are increasingly under pressure due to the rising prevalence of hearing loss worldwide (*World report on hearing* 2021; Haile et al., 2021). Reducing testing time whilst maintaining well-controlled sensitivity, preferably using existing clinical hardware, is therefore crucial to alleviate the burden on already overwhelmed health centres.

Here, we introduce and validate a new general-purpose framework for statistical inference in neurophysiological signals that explicitly accounts for the spectral colour and temporal dynamics of background noise. The proposed method, termed *Fmpi*, is a parametric method based on the well-established F-test (Elberling and Don, 1984; Don et al., 1984; Don and Elberling, 1994) which leverages information from within and across epochs to estimate the signal-to-noise ratio (SNR) and its statistical significance. Rather than relying on assumed or empirical population estimates of underlying distribution parameters, Fmpi derives analytical estimates of the effective degrees of freedom (DOF) directly from the power spectral density (PSD) of the recorded signal, thereby providing an individualised characterisation of statistical variability. By linking the spectral structure of the noise to the parameters governing the null distribution, this approach enables principled and computationally efficient inference without reliance on population-based assumptions or computationally intensive resampling procedures.

We demonstrate the utility of this framework in the context of AEP detection across multiple modalities, including ABRs, ASSRs, and cortical evoked potentials. Using both realistic simulations and large-scale human EEG datasets, we show that Fmpi achieves well-calibrated statistical performance under both stationary and non-stationary noise conditions, with sensitivity and specificity comparable to state-of-the-art bootstrapped methods, but at a fraction of the computational cost. In addition, the framework naturally supports estimation of the expected number of observations required for detection, enabling a principled futility stopping criterion that can substantially reduce recording time in clinical settings.

Although developed and validated here for AEP detection, the proposed framework is broadly applicable to other inferential problems in neurophysiology where coloured noise and temporal dependencies play a critical role. By providing an analytical link between noise structure and statistical inference, this work offers a general approach for robust, individualised analysis of neurophysiological data.

## 2 Methods

### 2.1 Fmpi

Most EPs are estimated by repeatedly presenting the evoking stimulus (Figure 1a), thereby reducing the underlying BN contaminating each presentation, (*y*_*i*_ (*t*)), and preserving the deterministic components of the evoked response through averaging. Once the average response is obtained, it is critical to assess whether sufficient evidence exists to consider the averaged waveform a true EP, rather than a random realisation of the post-averaged residual noise (RN).

**Figure 1.**
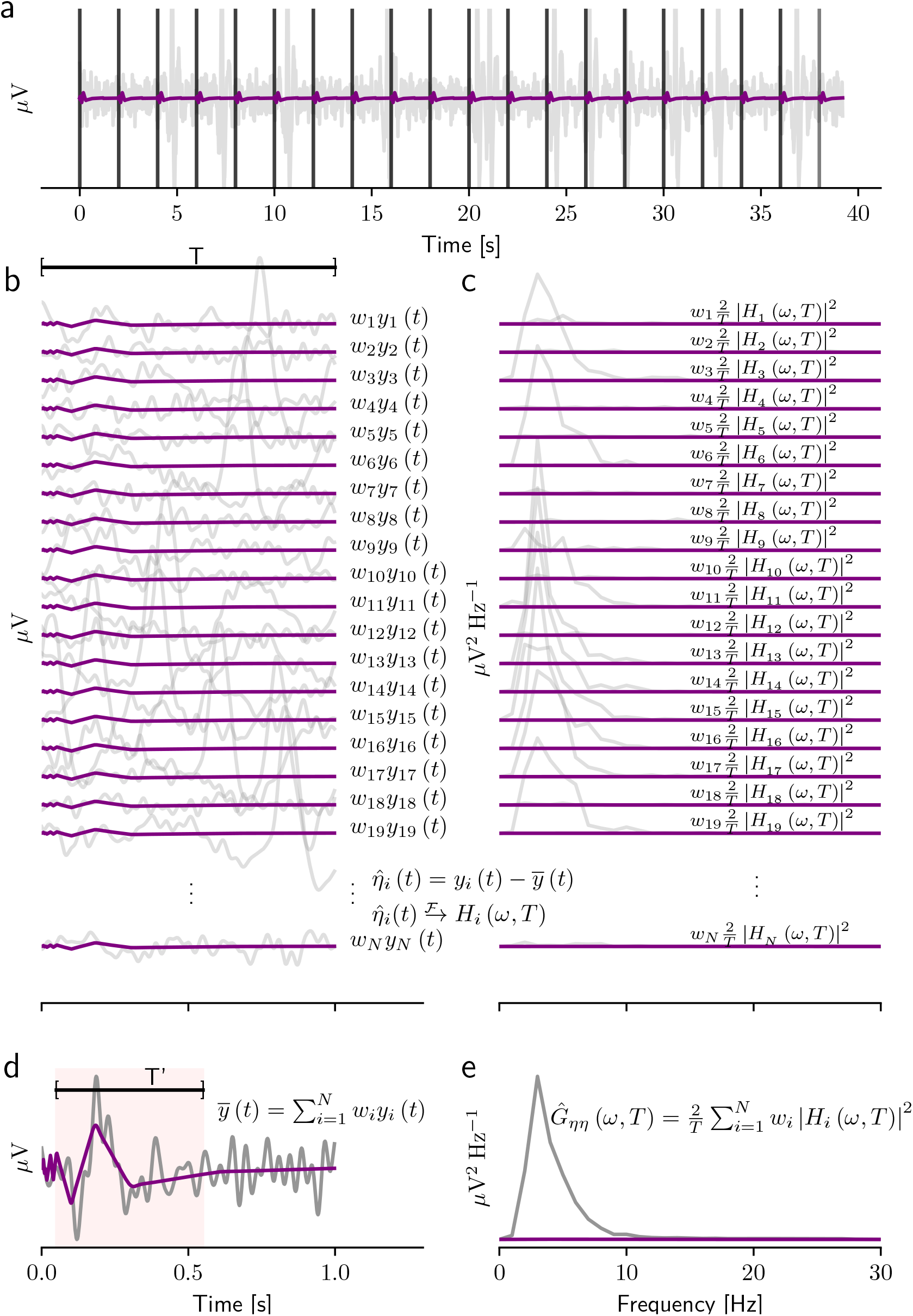
Evoked response estimation. The target EP is shown in purple. **a** Continuous time series consisting of the EPs and the recording BN is shown by the grey line. The timing of the event evoking the response is shown by the vertical black lines. **b** Single epochs (or epochs) of duration *T* (*y*_1_ (*t*), *y*_2_ (*t*),… *y*_*N*_ (*t*)) starting at the time of each event. Each of these epochs is assigned a weight (*w*_1_, *w*_2_,… *w*_*N*_). **c** Power spectral density (PSD)— 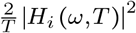 —of the estimated noise (*η*_*i*_ (*t*)) at each epoch. **d** Weighted averaged response in the time domain, and **e** weighted average PSD of the noise in the frequency domain.

The Fmpi is a parametric test that, like previous methods (Elberling and Don, 1984; Don et al., 1984; Don and Elberling, 1994), measures the ratio between the estimated variance of the averaged EP, 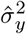, and the estimated variance of the residual noise, 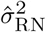, as shown in Eq. 1

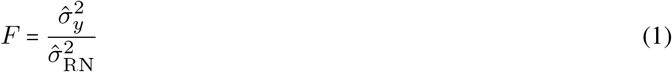

The probability that the F-value belongs to the null distribution, which represents the absence of a response, is given by Eq. 2

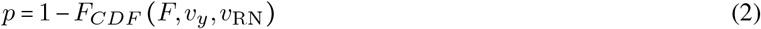

where *F*_*CDF*_ (*F, v*_*y*_, *v*_RN_) is the cumulative distribution function of an F-distributed random variable with DOF *v*_*y*_ and *v*_RN_.

Both *v*_*y*_ and *v*_RN_ are critical, as these parameters are used to set the threshold for objectively determining the presence or absence of an EP. The DOF are determined by the characteristics of the background noise, specifically its spectral properties and degree of stationarity, both of which are expected to exhibit variability across individual recordings. The Fmpi provides individualised estimates of *v*_*y*_ and *v*_RN_ for EPs analysed using either the time or the frequency domains, based on the properties—specifically, the spectral ‘colour’—of the EEG PSD. A detailed description and derivation of these estimates is provided in the Supplementary information.

The determination of the variances and their corresponding DOF depends on the analysis domain (time or frequency), as explained below.

#### 2.1.1. Time-domain responses

Time-domain EPs are typically measured by presenting repeated stimuli that evoke the response of interest (Figure 1a, purple trace). However, these recordings are contaminated by biological and endogenous noise (Figure 1a, grey trace). To mitigate this noise, the data are usually bandpass filtered to remove frequencies outside the EP bandwidth and segmented into epochs, *y*_*i*_ *t*, of constant duration *T*, covering the response time of interest. Noisy epochs may be excluded based on an empirical threshold or downweighted (Figure 1b) by applying per-epoch weights (*w*_*i*_, with ∑_*i*_ *w*_*i*_ = 1). To further reduce noise within the EP bandwidth (Figure 1b&c, grey traces) and approximate the true response (Figure 1 purple lines), the weighted epochs are averaged (Eq. 3; Figure 1d):

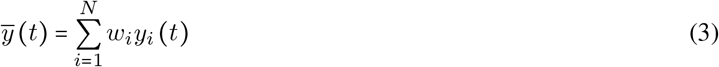

The post-average RN (see Supplementary information for details) can be estimated as:

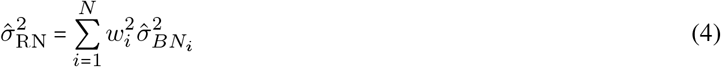

where the vector of weights is defined as:

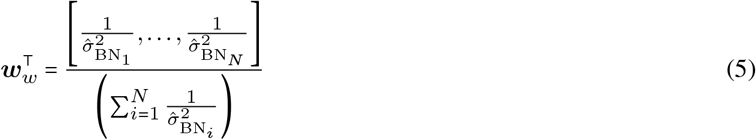

with 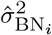 representing the per-epoch BN variance (described later) and with (·)^⊺^ denoting the transpose operator.

For time-domain EPs, it can be shown (Supplementary information) that the DOF of the RN variance (*v*_RN_), estimated from all available samples within each epoch of duration *T* are given by:

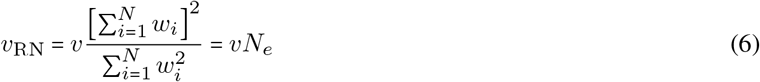

where *N*_*e*_ is the effective number of epochs estimated from the weights (*N*_*e*_ ≤ *N*) and *v* are the DOF within each epoch estimated from Eq. 7.

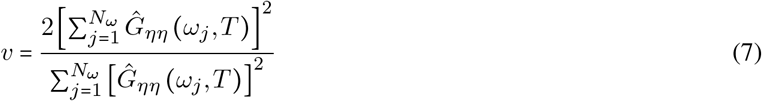

where 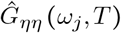 is the PSD (Figure 1e; Eq. 8):

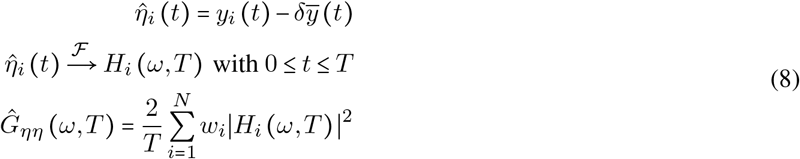

Here, *δ* (in this and subsequent equations) can be set to 0 if the power of the target response is much smaller than the background noise, or to 1 if the power of the target response is expected to be comparable to the background noise. Similarly, the DOF of the variance of the average response (*v*_*y*_) within a time window of interest *T* ^′^ ≤ *T* are given by:

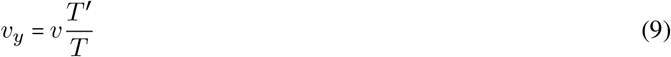

The variance of the BN 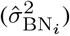, used to compute the weights, can be estimated either as the average variance of *P* fixed points across a number of epochs (*N*_*k*_), i.e., the across-epochs approach (Eq. 10; employed in the Fmpi-WAE approach), or from the variance within each epoch, i.e., the within-epochs approach (Eq. 11; employed in the Fmpi-WWE approach).

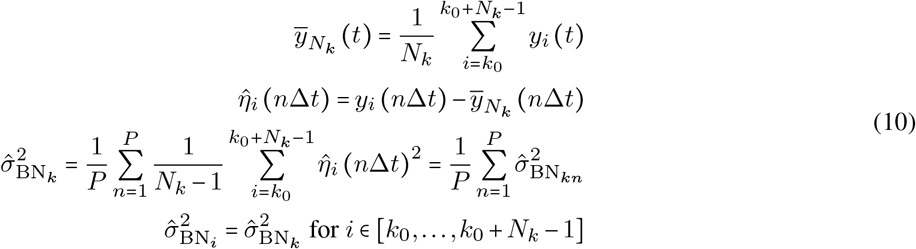

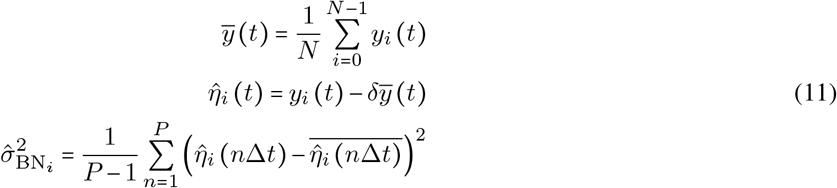

where *k*_0_ (Eq. 10) is the starting epoch of a block of *N*_*k*_ epochs, and Δ*t* is the time interval between the *P* tracked samples. In this study, P always corresponded to the number of samples available within an epoch, i.e., Δ*t =* 1/*f*_*s*_, with *f*_*s*_ being the sampling rate.

The choice of the noise variance estimation depends on the time scale of the response and the underlying noise dynamics. If the EP is measured using many (thousands of) epochs of short duration (<20 ms) at a high repetition rate (as in ABRs), where the dynamic fluctuations of the BN are slow relative to the repetition rate (e.g., slow myogenic artefacts lasting hundreds of milliseconds), the across-epochs approach (Eq. 10) is more accurate. This is because the total number of data points used to estimate the common noise variance within a block of epochs is greater than if only the data of a single epoch were used, thereby better capturing the dynamic fluctuations of the BN for weight estimation. In contrast, for cortical EPs, which typically consist of a few hundred epochs of long duration (>800 ms) obtained at a low repetition rate, the variance of the BN can be accurately estimated within each epoch.

#### 2.1.2. Frequency-domain responses

Several types of physiological responses (e.g., ASSR, frequency following responses (FFRs), steady-state visual evoked potentials (SSVEPs), and otoacoustic emissions (OAEs)) evoke frequency-specific activity. In such cases, it is necessary to determine whether the power of the frequency components of interest is significantly greater than the power of the underlying noise. The Fmpi can be conveniently applied for this purpose. Two approaches are provided: one which is suitable for narrow-to-wide band frequency analysis, and another suitable for sparse or single-frequency component analysis.

#### 2.1.3. Fmpi in the frequency-domain—narrow-band analysis

Consider a narrow-band, weighted-averaged signal (Eq. 3) with frequencies of interest *ω*_*j*_ ∈ [*w*_*a*_, *w*_*b*_, …, *w*_*m*_] . The post-average variance (Supplementary information) is given by Eq. 12, whilst the variance of the BN within each epoch is given by Eq. 13.

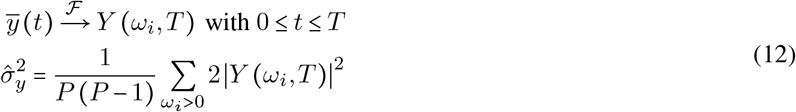

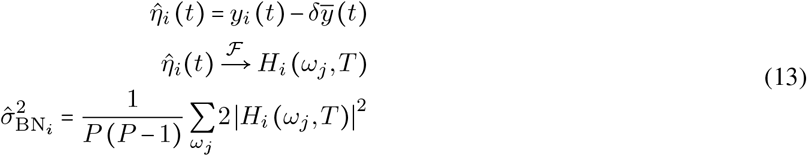

The frequency-specific residual noise is obtained via Eq. 4 and the F-value via Eq. 1.

The DOF of the F-value are obtained from the weighted-averaged PSD (Eq. 8) as shown below:

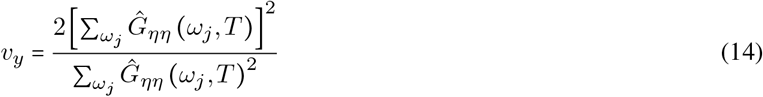

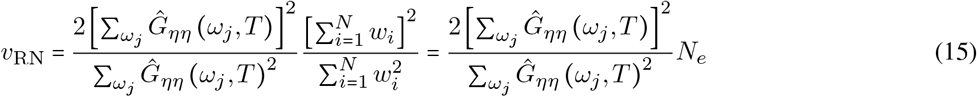

#### 2.1.4. Fmpi in the frequency-domain—sparse-frequency analysis

In cases where only a few M sparsely distributed frequencies of interest *ω*_*j*_ ∈ [*w*_*a*_, *w*_*b*_, …, *w*_*m*_], such as the fundamental frequency and several harmonics, are relevant, it is necessary to assess whether the pooled power of these frequencies is significantly greater than the power of the underlying background noise at the same frequencies. This can be achieved by assuming that the noise at the frequencies of interest is similar to that at their neighbouring frequencies. Under this assumption, the pooled variance of the BN can be more accurately estimated—with increased DOF—by averaging the power of 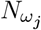 frequencies in the neighbourhood of each frequency of interest (Eq. 16).

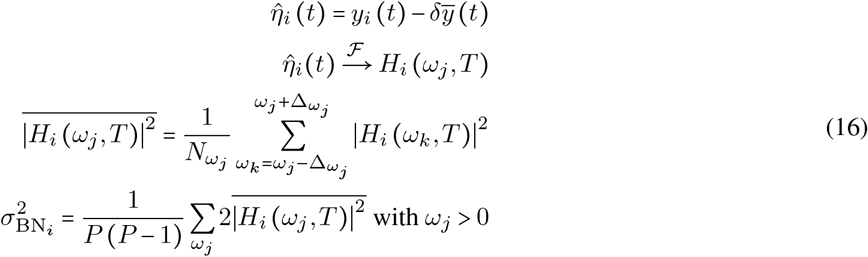

The frequency-specific residual noise variance is obtained using Eq. 4, the weighted-averaged signal variance using Eq. 12, the F-value using Eq. 1, the DOF of the weighted-averaged signal using Eq. 14, and the DOF of the post-average RN is given by Eq. 17.

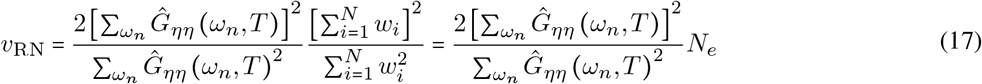

with *ω*_*n*_ ∈ [*ω*_1_ − Δ*ω*_1_, …, *ω*_1_ + Δ*ω*_1_, *ω*_2_ − Δ*ω*_2_, …, *ω*_2_ + Δ*ω*_2_, …, *ω*_*m*_ − Δ*ω*_*m*_, …, *ω*_*m*_ + Δ*ω*_*m*_].

#### 2.1.5. Futility

In many situations, it is desirable to estimate how much longer a measurement should continue—either to predict the total recording time required to detect an EP, or, alternatively, to terminate the recording earlier if there is insufficient evidence that an EP will be detected within a reasonable timeframe (i.e., futility stopping). The Fmpi can be used to predict and update the necessary number of epochs (*N*_*s*_), based on the ongoing information obtained from N epochs during a recording. This estimation is given by Eq. 18 (see Sup. 1.12.4 for details).

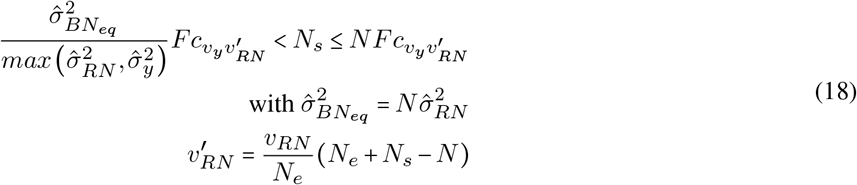

Here, *N*_*s*_ is the predicted number of epochs required to detect a response, based on the information obtained from the current *N* epochs—specifically, the current 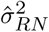, the current DOF (*v*_*y*_ and *v*_*RN*_), and the current effective number of epochs *N*_*e*_. *Fc* is the critical F-value required to reject the null-hypothesis.

The right side on Eq. 18 reflects the upper limit imposed by the fact the variance of the averaged response 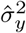 cannot be smaller than the variance of the residual noise 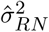 (see Sup. 1.12.4 for details). This relationship can be exploited to estimate estimate the DOF of the residual noise (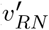 in Eq. 18) at the predefined maximum number of epochs *N*_*f*_ —the futility threshold—and, consequently, to determine the critical F-value (*Fc*) required to reject the null hypothesis. Importantly, the critical F-value depends on both *v*_*y*_ and 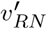.

If the futility threshold is unknown, an alternative approach is to estimate a threshold that ensures a clinically acceptable RN with variance 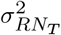, using Eq. 19.

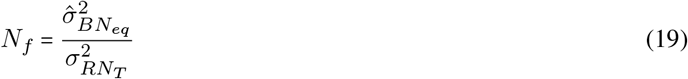

Under the assumption that recording conditions remain constant, this framework enables prediction of whether the number of epochs required to obtain a statistically significant response will exceed the futility threshold. Measurements can therefore be terminated once the condition *N*_*s*_ > *N*_*f*_ is met.

Notably, in the absence of a neural response, *N*_*s*_ *= NFc*; that is, the number of epochs required to detect a response will increase by a factor of *NFc*, where the critical F-value is determined by the spectral characteristics—colour—of the individual underlying noise.

### 2.2. Fmp statistics

The original Fmp method (Elberling and Don, 1984; Don et al., 1984; Don and Elberling, 1994) employing weighted averaging across epochs (Fmp-WAE) utilised an F-value calculation that was identical to that of the Fmpi-WAE. The sole distinction was that, in the original Fmp-WAE, the degrees of freedom (DOF) were fixed at *v*_*y*_ *=* 5 and *v*_*RN*_ *= N* (Elberling and Don, 1984; Don et al., 1984; Don and Elberling, 1994). For ASSRs, the Fmp-WAE was computed in the same manner as the frequency-based Fmpi-WAE; however, the degrees of freedom were unweighted, specifically *v*_*y*_ *=* 2*N*_*ω*_ and *v*_*RN*_ = *N*, where *N*_*ω*_ denotes the number of frequency bins included in the analysis.

### 2.3. Hotelling-T2 and modified T2 statistics

The Hotelling-T2 (Hotelling, 1931) is a multivariate method that has been widely employed for the detection of EPs in both the time and frequency domains (Picton et al., 2003; Golding et al., 2009; Chesnaye et al., 2018; Bardy et al., 2020; Chesnaye et al., 2021a; Chesnaye et al., 2023). The Hotelling-T2 statistic is defined by the following equations:

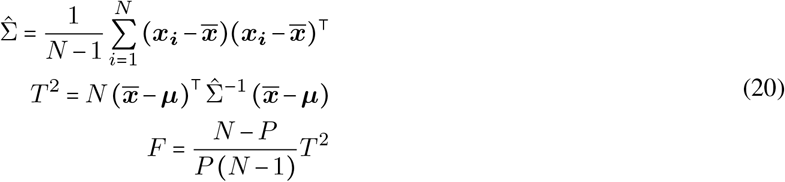

Here, F follows a Fisher–Snedecor distribution with DOF *P* and *N* − *P*, where *N* is the number of epochs and *P* the number of features contained in the mean feature vector 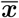. The mean feature vector 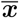 is compared with 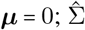 is the sample covariance matrix, and ⊺ denotes the transpose operator. The *P* mean features in 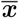 correspond to the averages of across the N columns of the feature matrix ***x***_*P* ×*N*_.

In this study, the feature matrix was constructed using attributes computed either in the time domain (i.e., several amplitude means within a predefined analysis time window) or in the frequency domain (i.e., real and imaginary Fourier coefficients from a Fourier transform applied to the analysis time window) (Golding et al., 2009; Chesnaye et al., 2018; Bardy et al., 2020; Chesnaye et al., 2021a; Chesnaye et al., 2023). To distinguish between these approaches, the acronym HT2 was used for time-domain-based features, and HT2-f for frequency-domain-based features. For frequency-domain-based features, either the full sample covariance matrix was used (HT2-f), or a version in which all off-diagonal elements were set to zero (HT2-f-diag). For the latter, the null distribution was estimated either under the assumption that the DOF are the same as with the full sample covariance matrix, or empirically by bootstrapping the *T* ^2^ statistic from Eq. 20 (see Bootstrapping)—the modified T2 statistics test (T2-f-diag; Chesnaye et al., 2023).

It should be noted that 1) the F-value in Eq. 20 depends on both the number of epochs (*N*) and the number of features (*P*), 2) that the statistical power of the Hotelling-T2 decreases as the number of features increases—the so-called ‘small n large p’ problem (Bai and Saranadasa, 1996)—, and 3) that the method becomes invalid when *P* >= *N*.

### 2.4. Q-sample

In addition to Fisher–Snedecor based detection methods, we employed the modified Q-sample test (Stürzebecher et al., 1999; Cebulla et al., 2006; Stürzebecher and Cebulla, 2013; Cebulla and Stürzebecher, 2015; Chesnaye et al., 2021a; Chesnaye et al., 2023). The Q-sample test is a non-parametric approach that calculates the *W* test statistic from *q* frequency bins obtained for each of the *N* epochs, as shown below:

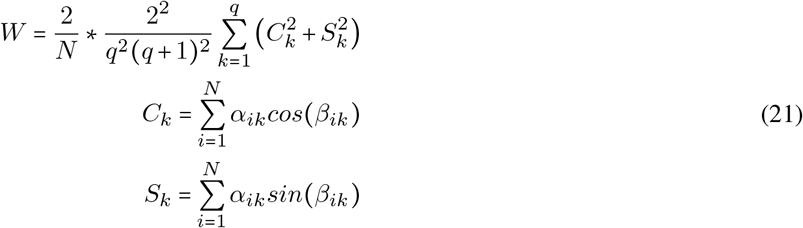

Here, *α*_*ik*_ may represent either the original spectral amplitude of the frequency bin or a ranked version thereof, and *β*_*ik*_ may be either the original spectral phase or a ranked version. In this study, the original phase was retained, and the amplitude of the frequency bins was ranked uniformly within the range *α*_*ik*_ ∈ 1, 2, 3, …, *Nq* (Chesnaye et al., 2021a; Chesnaye et al., 2023). The ranking strategy employed in the Q-Sample test aims to avoid excessive weights to epochs with large artefacts, especially when the number of epochs is reduced. The null distribution was obtained empirically via bootstrapping (see Bootstrapping).

### 2.5. Simulations

To evaluate the performance of the proposed Fmpi method, we conducted realistic simulations of EPs. These simulations utilised a template response (Figure 2a, target) embedded in 1/*f* pink noise, which closely approximates human brain background activity (He et al., 2010), under both stationary and non-stationary noise conditions (the latter including eye-blinking artefacts). Pink noise was generated by filtering a Gaussian white noise signal using a finite impulse response (FIR) filter with a 1/*f* ^2^ transfer function. Eye-blink artefacts were simulated by randomly assigning unitary impulses over time at an average interval of 2 s (SD = 1.4 s). These impulses were subsequently convolved with a human eye-blink template (Valderrama et al., 2018) to generate the artefact waveform.

**Figure 2.**
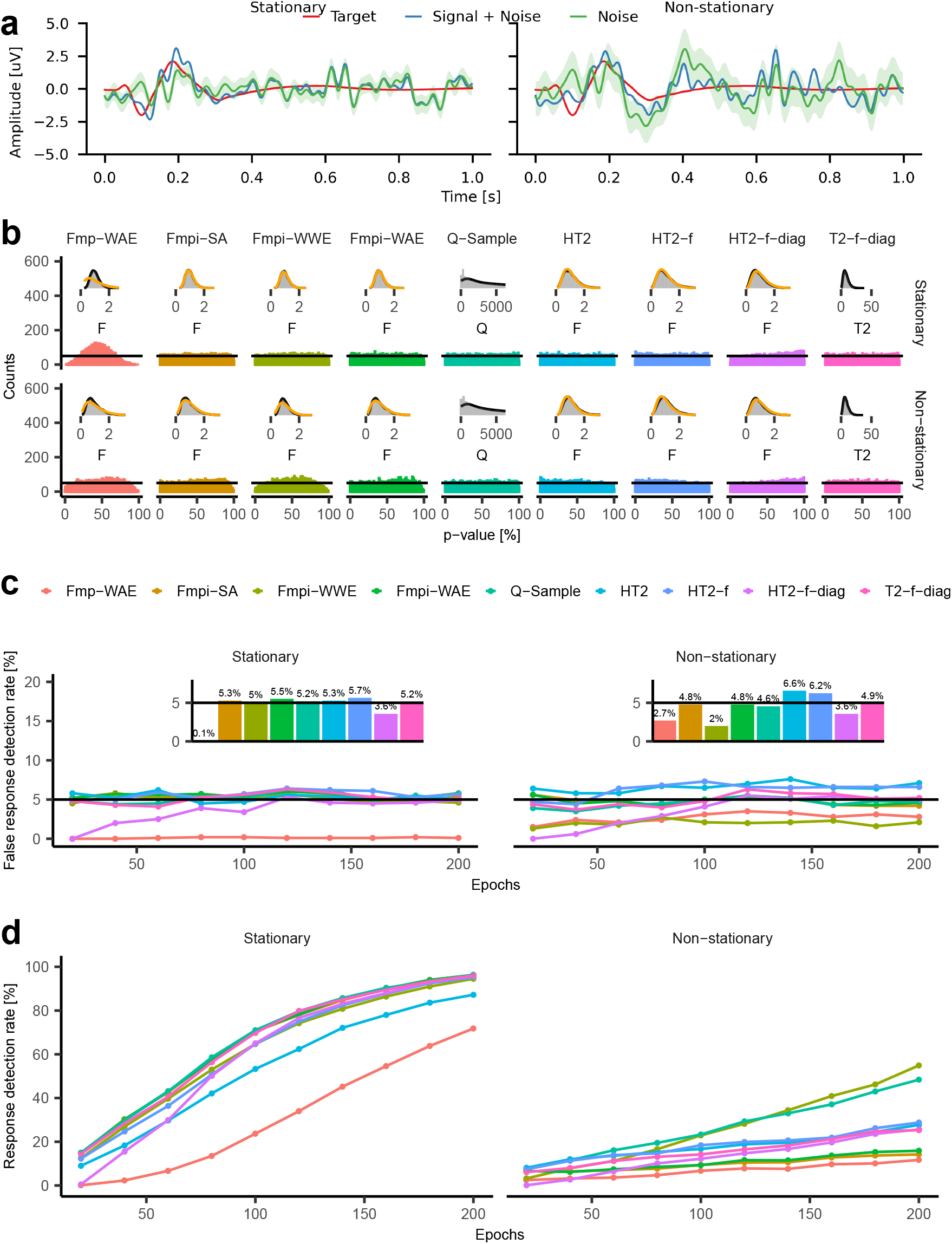
Fmpi exhibits robust sensitivity and specificity over time under stationary and non-stationary noise in cortical response simulations. **a** Examples of simulations under stationary (left) and non-stationary (right) background noise. The template cortical response (Target) is shown by red lines. Weighted average waveforms containing the cortical template are shown by light blue lines (Signal + Noise), whilst averages obtained without cortical responses (Noise) are shown in green, with the standard error of the mean indicated by the shaded green area. **b** P-value histograms (200 bins) obtained in the absence of cortical responses for stationary (top) and non-stationary (bottom) noise. The horizontal solid black lines represent the ideal unbiased distribution. Insets show the empirical distribution of the F-, Q-, or T2-values versus the theoretical (orange line) PDFs of an *F* -distribution pooled across epochs. The DOF, *v*_1_ and *v*_2_, correspond to the median of the pooled DOF estimated for each method at each epoch block. **c** False response detection rates (noise-only data) obtained at a nominal sensitivity of 95%. Insets show false response detection rates pooled across all epochs. **d** response detection rates in the presence of a cortical response (Signal + Noise) as a function of the number of epochs. A total of 1000 simulations were employed in each epoch. Left panels show stationary noise; right panels show non-stationary noise. The number of simulations per epoch block is indicated by the lines connected to the data points.

For cortical simulations, the power of the stationary background noise was adjusted such that the final SNR within the analysis filter bandwidth (2-30 Hz) was 3 dB after 200 epochs (Figure 2a, Signal + Noise). For ASSR simulations, the same template signal was convolved with a train of unitary impulses presented at a rate of 41 Hz, producing an ASSR with energy at 41 Hz and its harmonics (Sup. Figure 1). Each simulation comprised 80 epochs of 1 second duration, resulting in a final SNR of ≈ 12 dB under stationary noise. Simulations were carried out twice: once with the neural response included, and once without, using identical noise in both cases (Figure 2a, Noise).

### 2.6. Recordings

To test the Fmpi detector, we analysed data from multiple studies, encompassing ABRs, ASSRs, and cortical recordings. These datasets included both research and clinical recordings from adults with normal and impaired hearing, as well as from infants. Detailed descriptions of the datasets are provided in Table 1.

**Table 1.** Summary of the data and EPs analysed in this study. Columns from left to right: study ID; data source; EP type; stimulus; number of participants (N); total number of measurements (M); number of EEG channels included (C); age, hearing status (NH or HI); ethical approval; and EEG recording system. The studies encompass cortical ACC responses to interaural phase differences (IPDs), interaural time differences (ITDs), frequency, or spectrotemporal transitions; cortical AEPs to speech sounds; ASSRs to various amplitude modulations (AMs) and binaural stimuli (IPM-FRs and ITD-FRs; and ABRs to narrow-band CE-chirp, tone-bursts, and clicks.

| Study | Source | Type | Stimulus | N | M | C | Age | Hearing | Ethics | EEG system |
| --- | --- | --- | --- | --- | --- | --- | --- | --- | --- | --- |
| 1 | Van Yper et al. (2023) | Cortical ACC | IPM; -90/90° & 0/180° IPD changes; 40 and 80-Hz AMs; test-retest | 30 | 240 | 2 | 18-42 years | NH | Macquarie University ethics committee (52021646228840) | BioSemi ActiveTwo (Amsterdam, The Netherlands) |
| 2 | Simonsen et al. (2025) | Cortical ACC | Spectrotemporal modulations; 0-to-16 dB contrast levels | 16 | 40 | 2 | 20-79 years | NH & HI (audibility compensated) | Science-Ethics Committee for the Capital Region of Denmark (H-1-2013-138) | Modified Eclipse system (Interacoustics, Middelfart, Denmark) |
| 3 | Rekswinkel et al. (2023) | Cortical ACC | Spectral changes; vowel-like and pure tones stimuli (Martin and Boothroyd, 2000); ACC responses to frequency and/or amplitude changes | 45 | 182 | 2 | 21-81 years | NH & HI (audibility compensated) | Science-Ethics Committee for the Capital Region of Denmark (H-1-2013-138) | BioSemi ActiveTwo (Amsterdam, The Netherlands) |
| 4 | Chesnaye et al. (2023) and Visram et al. (2023) | Cortical AEP | Synthetic speech tokens (Stone et al., 2019); mid and high frequency; 0 to 30 dB SL | 48 | 188 | 1 | 3-7 months | HI (aided) | North West National Research Ethics Service Ethics Committee (15/NW/0736) | Modified Eclipse system (Interacoustics, Middelfart, Denmark) |
| 5 | Kristensen et al. (2026) | ABR | NB CE-Chirp (Elberling and Don, 2010); left & right ears; 0.5, 1, 2, & 4 kHz; 35-85 dB nHL; consecutive or simultaneous presentation; test & retest | 25 | 2785 | 2 | 20-29 years | NH | Science-Ethics Committee for the Capital Region of Denmark (H-1-2013-138) | Modified Eclipse system (Interacoustics, Middelfart, Denmark) |
| 6 | Lightfoot and Stevens (2013) | ABR | 4kHz tone burst; 30 to 85 dB nHL | 25 | 240 | 1 | 1-12 weeks | Referred from the English newborn screen | Arrowe Park Hospital, Wirral, United Kingdom | Modified Eclipse system (Interacoustics, Middelfart, Denmark) |
| 7 | Bowen et al. (2018) | ABR | 100 μs clicks; 60 peSPL; diotic stimulation | 20 | 20 | 2 | 19-42 years | NH | Macquarie University ethics committee (5201600895) | BioSemi ActiveTwo (Amsterdam, The Netherlands) |
| 8a | Van Yper et al. (2023) | ASSR | 81-Hz AMs under -90/90° & 0/180° IPD changes; test-retest | 30 | 120 | 2 | 18-42 years | NH | Macquarie University ethics committee (52021646228840) | BioSemi ActiveTwo (Amsterdam, The Netherlands) |
| 8b | Van Yper et al. (2023) | ASSR | 40-Hz AMs under -90/90° & 0/180° IPD changes; test-retest | 30 | 120 | 2 | 18-42 years | NH | Macquarie University ethics committee (52021646228840) | BioSemi ActiveTwo (Amsterdam, The Netherlands) |
| 8c | Van Yper et al. (2023) | ASSR | 6.7-Hz IPM-FRs; -90/90° & 0/180° IPD transitions; 40 and 81-Hz AMs; test & retest | 30 | 240 | 2 | 18-42 years | NH | Macquarie University ethics committee (52021646228840) | BioSemi ActiveTwo (Amsterdam, The Netherlands) |
| 9a | Undurraga et al. (2020b) | ASSR | 81-Hz AMs under 6 ITDs | 23 | 138 | 2 | 18-35 years | NH | Macquarie University ethics committee (52020527017402) | BioSemi ActiveTwo (Amsterdam, The Netherlands) |
| 9b | Undurraga et al. (2020b) | ASSR | 6.7-Hz ITD-FRs; 4 ITDs; 81-Hz AM | 23 | 92 | 2 | 18-35 years | NH | Macquarie University ethics committee (52020527017402) | BioSemi ActiveTwo (Amsterdam, The Netherlands) |
| 10 | Visram et al. (in preparation) | ASSR | 38.1, 68.4, and 69.3-Hz simultaneous AMs for 0.7, 2, and 4-kHz carriers, respectively, using speech-like NB CE-Chirp (Laugesen et al., 2018) | 40 | 375 | 2 | 3-7 months | HI (aided) | North West National Research Ethics Service Ethics Committee (15/NW/0736). | Modified Eclipse system (Interacoustics, Middelfart, Denmark) |

### 2.7. Processing

To compare the performance of all detectors, identical datasets were processed using each detection method. These included the Fmpi with standard average (Fmpi-SA; unweighted average), Fmpi with within-epochs (Fmpi-WWE) or across-epochs (Fmpi-WAE) weighted averaging, Fmp-WAE (Don and Elberling, 1994; Stürzebecher et al., 1999; Cebulla et al., 2006; Silva, 2009; Golding et al., 2009; Stürzebecher and Cebulla, 2013; Cebulla and Stürzebecher, 2015; Chesnaye et al., 2018; Bardy et al., 2020; Chesnaye et al., 2021a; Chesnaye et al., 2023), Hotelling-T2 test using features from both the time (HT2) and frequency (HT2-f and HT2-f-diag) domains, and the fully bootstrapped T2-f-diag (Chesnaye et al., 2023) and Q-Sample methods.

For ABR analyses, all data were downsampled to 8000 Hz, with epochs of 20 ms duration and an analysis window of 15 ms (either 2-17 ms or 5-20 ms, depending on stimulus characteristics). Cortical recordings were downsampled to 500 Hz, with epochs of ≈ 1 s and an analysis window of 500 ms (50 to 550 ms).

ABR data were band-pass filtered between 68 and 1350 Hz, and cortical responses between ≈ 2 and 30 Hz (Kaiser linear phase filter, 65 dB stop-band attenuation, 1 Hz transition between pass and stop bands). The high-pass cutoff frequency was set to the inverse of the analysis window length to reduce bias in F-value estimation for the Fmp-WAE and Fmpi methods (Elberling and Don, 1984; Don and Elberling, 1994; McKearney et al., 2023).

To capture the temporal dynamics of each method as a function of available data, recordings were analysed in incremental blocks of epochs: 200 for ABR, 20 for cortical, and 5 for ASSR recordings. To maximise the number of independent measurements per block, all possible independent blocks within each recording were included. For example, an ABR recording with 4000 epochs yielded 20 independent blocks of 200 epochs, 10 blocks of 400 epochs, 6 blocks of 600 epochs, 5 blocks of 800 epochs, and so forth. Due to the high computational cost of bootstrapping with ABR recordings, the number of included epochs was limited to 4000.

Block sizes for weight estimation with Fmp-WAE and Fmpi-WAE were 200, 20, 5 and epochs for ABR, cortical, and ASSR measurements, respectively. The *δ* parameter in equations Eq. 8, Eq. 11, Eq. 13, and Eq. 16 was set to zero, as the energy of the BN was typically more than 20 dB greater than that of the EPs analysed in this study.

To evaluate the accuracy and robustness of the detection methods under conditions of non-stationary noise, the rejection of noisy epochs was deliberately omitted.

#### 2.7.1. Feature selection

Time-domain detectors utilised various features, all extracted within a consistent analysis window. For the HT2 method, amplitude means served as features, comprising either 40 (375 µs) amplitude means for ABRs (Chesnaye et al., 2018), or 10 (50 ms) amplitude means for cortical analyses (Golding et al., 2009; Bardy et al., 2020; Chesnaye et al., 2021a). For cortical responses, frequency-domain detectors employed frequency bins between ≈ 2 and 8 Hz (yielding 4 bins or 8 features; Chesnaye et al., 2023), and between ≈ 67 and 600 Hz (9 bins or 18 features) for ABRs.

For ASSRs, all detectors incorporated the same features: the real and imaginary components of the frequencies of interest (i.e., the evoking frequency and its harmonics). In ASSR simulations and in all studies except one (study 10, Table 1) three harmonics (six features) were used—the fundamental and the next two harmonics. In study 10, twelve harmonics (24 features) were employed (Cebulla et al., 2006; Stürzebecher and Cebulla, 2013; Cebulla and Stürzebecher, 2015). The choice between three or twelve harmonics was guided by two considerations. First, as indicated before, the performance of the Hotelling-T2 can be compromised as the number of features increases (‘small n large p’ problem) (Bai and Saranadasa, 1996). Second, the averaged ASSR waveforms from studies 8 and 9 (Figure 7a and Figure 8a) exhibited prominent peaks primarily at the first three harmonics. Thus, using three harmonics was intended to preserve good performance for HT2-f and HT2-f-diag while retaining relevant information. However, since previous studies employing the Q-Sample method included between six and twelve harmonics, twelve harmonics were used in study 10—accepting a reduction in performance for frequency-based Hotelling-T2 methods—to maintain comparability with Q-Sample analyses (Stürzebecher and Cebulla, 2013; Cebulla and Stürzebecher, 2015).

The bandwidth for noise estimation in the frequency-domain Fmpi method was set to 4 Hz (Δ*ω*_*j*_ *=* 2 Hz in Eq. 16).

#### 2.7.2. No response data

Due to the limited availability of ABR and cortical recordings obtained without stimulation—which are essential for assessing the sensitivity of different detection methods—we generated no-response datasets from the original recordings. First, the evoked response was estimated for each recording (Figure 1d) using all available epochs. This estimated response was then subtracted from the raw data (Figure 1a), at each event onset. Subsequently, the original event times were replaced by random event times (within the time range of the data) to ensure that any residual evoked response was eliminated when averaging the epochs (i.e., incoherent averaging). This procedure also prevented the final average from converging to zero as the number of epochs approached the total used to generate the subtracted evoked response. Simulations and analysis of real data (not shown) confirmed that this approach preserved the properties of the underlying noise—such as power, spectral shape, and correlation—whilst effectively removing the evoked response. For ASSRs, the evoked response was not subtracted; instead, analyses were performed on neighbouring frequency bins where no response was expected.

#### 2.7.3. Bootstrapping

As previously described, we utilised the non-parametric Q-sample (Stürzebecher et al., 1999; Cebulla et al., 2006; Stürzebecher and Cebulla, 2013; Cebulla and Stürzebecher, 2015; Chesnaye et al., 2021a) and the modified T2 statistic test (Chesnaye et al., 2023), with null distributions estimated empirically via bootstrapping (Efron and Tibshirani, 1993; Lv et al., 2007). First, the evoked response was subtracted from the continuous data (as outlined above). Subsequently, new random event times—equal in number to the original events and uniformly distributed within the original event time window—were generated. These randomised event times were applied to the continuous data, and either the *W* or T2 statistics were calculated from the resulting set of surrogate epochs. For each tested point within each recording, a total of 1000 test values were bootstrapped to construct the null distribution (Chesnaye et al., 2021a).

#### 2.7.4. Performance metrics

Detector performance was evaluated using several metrics, including detection bias, false response detection rate, response detection rate, receiver operating characteristic curve (ROC), area under the curve (AUC), and average area under the curve (AAUC). Detection bias was quantified from the p-value distribution obtained from the no-response datasets (Figure 2b), which should be uniformly distributed for an unbiased detector (Murdoch et al., 2008). Observed sensitivity was calculated by integrating the p-value distribution (Figure 3a) and bias was defined as the mean absolute difference between nominal and observed sensitivity (Figure 2b), assessed either across the full sensitivity range (0-100%) or within the clinically relevant range (95-100%). False response detection rates (from no-response data) and response detection rates (from original datasets) were determined as a function of time (in epochs) at a nominal sensitivity of 95% (Figure 2c&d), by pooling all available detections for each participant within each study.

**Figure 3.**
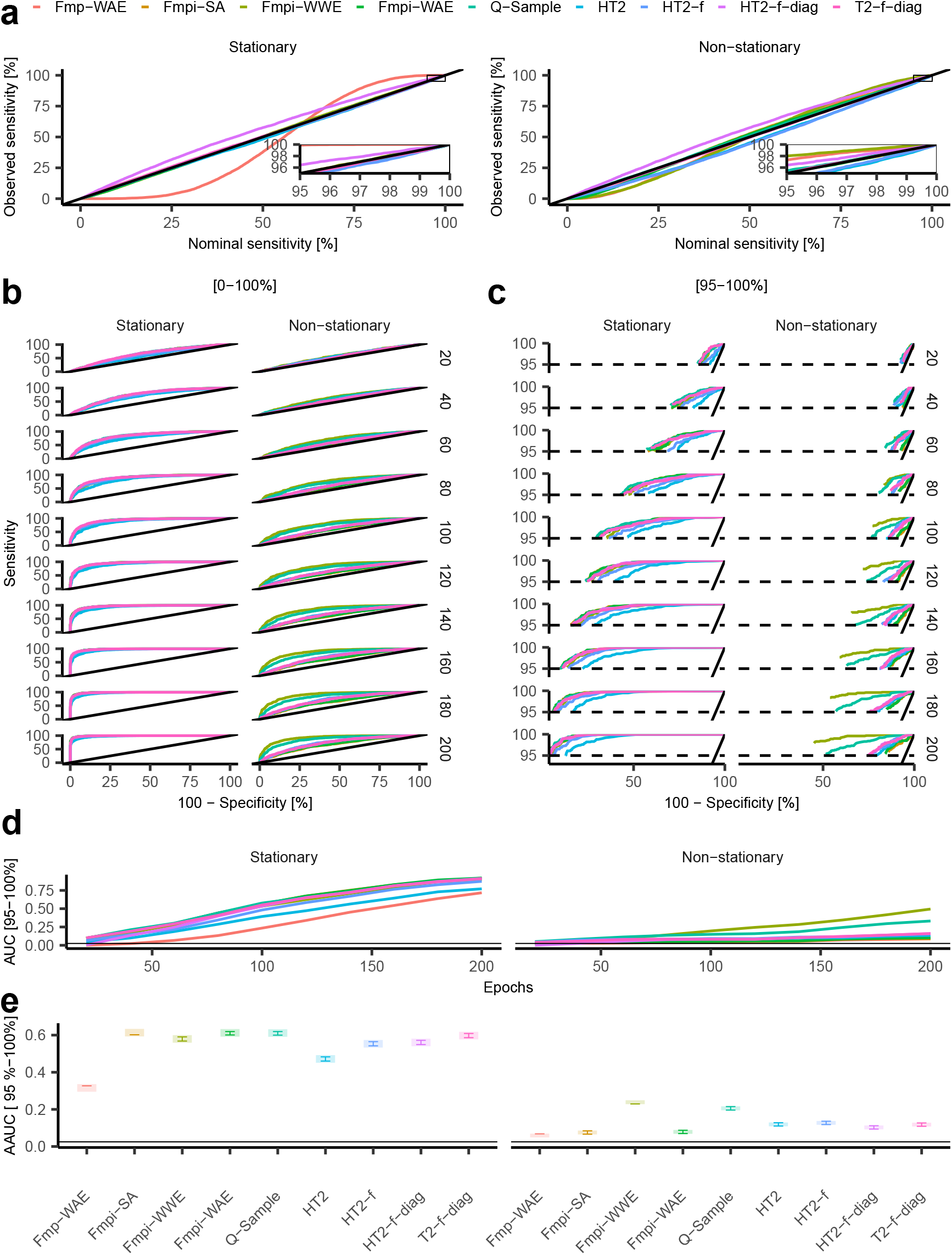
Fmpi enables high performance across the span of clinically relevant sensitivity rates in simulated cortical responses. **a** Nominal versus observed sensitivity for stationary (left) and non-stationary (right) noise across all possible sensitivities. Traces represent data pooled across all epochs. The diagonal solid black line indicates the ideal unbiased expectation. Insets provide a zoomed view of the clinically relevant sensitivity regions (95-100%). **b** ROCs for stationary (left) and non-stationary (right) noise for all possible sensitivities at different number of averaged epochs (indicated on the right side of the panels). The diagonal solid black line represent the chance level of the ROCs. **c** Same as **b**, but computed for the clinically relevant nominal sensitivities (95-100%). The 95% nominal sensitivity is indicated by horizontal dashed lines. **d** AUC as a function of the number of epochs for the clinically relevant nominal sensitivities shown in **c**. Note that the AUCs have been normalised by the total area within the clinically relevant region. The corresponding chance level is shown by the horizontal solid black line. **e** AAUC for the clinically relevant nominal sensitivities. Shaded areas represent 95% confidence intervals. The length of the error bars has been adjusted to allow for visual post hoc comparisons (Holm FWER corrected). Non-overlapping segments indicate significant differences, whilst overlapping segments indicate non-significantly differences. The horizontal solid black line indicates the chance level of the AAUC.

ROCs were constructed for different testing times (in epochs; Figure 3b&c) using sensitivity and specificity rates obtained for all possible nominal sensitivities or specifically for the clinically relevant range (95-100%). The AUC as a function of time (Figure 3d) was calculated as the area under each ROC and normalised by the total possible area, i.e., by 1 for all possible nominal sensitivities—with a corresponding chance level of 0.5—or by 0.05 for the clinically relevant range of nominal sensitivities (95-100%)—with a corresponding chance level of 0.025. Finally, the AAUC (Figure 3e), representing the overall performance of each detector, was estimated by averaging the AUC across all epochs.

All EEG simulations and recordings were processed offline using pEEGy v2.0.0, an open-source Python 3–based analysis package (Undurraga, 2026).

### 2.8. Statistical analysis

All statistical analyses were conducted using linear mixed-effects (LME) models (Bates et al., 2015). In all models, the dependent variable was the AAUC, the independent factor was the *detector* type, and the random factor was the *subject*. For the simulations, the 1000 runs were grouped into blocks of 20, with these groups treated as the random factor in the models. The DOF were estimated using the Satterthwaite approximation (Kuznetsova et al., 2017). Conditional, random, and marginal residuals of the LME models were inspected visually (Lüdecke et al., 2021).

When outliers were identified (typically fewer than 2% based on the 1.5 inter-quartile range criterion), they were removed and the LME model was refitted. Conditional, random, and marginal residuals of the models were checked by visual inspection. Most outliers originated from the Fmp-WAE and HT2-f-diag detectors, which exhibited lower performance compared to other detectors. Post hoc comparisons were performed using Holm’s family-wise error rate (FWER) correction (Russell V. Lenth, 2024).

All statistical analyses were performed using the R software package (R Development Core Team, 2024), with the significance level set at *α =* 0.05.

## 3. Results

We implemented three versions of the Fmpi algorithm, each differing in their approach to handling temporally non-stationary noise through distinct temporal weighting strategies (see Methods). These strategies include: (1) standard average (Fmpi-SA), where all epochs are weighted equally; (2) weighted average within epoch (Fmpi-WWE), where each epoch is weighted based on its internal variance; and (3) weighted average across epochs (Fmpi-WAE), where subsets of consecutive epochs share the same weights. We compared these Fmpi variants with several established parametric algorithms, including the clinical established time-based weighted average method (Fmp-WAE; Elberling and Don, 1984; Don et al., 1984; Don and Elberling, 1994), a time-based Hotelling-T2 (HT2; Golding et al., 2009; Chesnaye et al., 2018; Bardy et al., 2020; Chesnaye et al., 2021a)—a multivariate generalisation of Student’s t-statistic—, two frequency-based Hotelling-T2 (HT2-f and HT2-f-diag; Chesnaye et al., 2023), and two frequency-based, fully bootstrapped methods: the non-parametric Q-Sample test (Stürzebecher et al., 1999; Cebulla et al., 2006; Stürzebecher and Cebulla, 2013; Cebulla and Stürzebecher, 2015; Chesnaye et al., 2021a) and the modified T2 statistics test (T2-f-diag; Chesnaye et al., 2023).

The primary distinction between the Fmpi and Hotelling-T2 variants lies in the estimation of the DOF. The DOF (*v*_1_ and *v*_2_) are the two parameters that determine the EP response detection threshold from the Fisher–Snedecor probability distribution—F-distribution—assumed by both methods. The Fmpi estimates both DOF based on the individual spectral characteristics—i.e., the ‘colour’—of the background noise and its non-stationary temporal fluctuations (see Methods). In contrast, Hotelling-T2 based methods, although capable of accounting for individual temporal correlations in the data, rely on fixed DOF determined by the dimensionality of the selected feature set (e.g., mean amplitudes from multiple time windows or frequency components) and the number of epochs.

The Fmp-WAE method, which also assumes an F-distribution, sets one DOF (*v*_1_) to a fixed value, derived empirically from human EEG recordings (Elberling and Don, 1984; Don et al., 1984; Don and Elberling, 1994). The Q-Sample method transforms frequency-based features into ranks to reduce the effects of large non-stationary noise events. The response detection threshold is set according to the empirical properties of the underlying noise, estimated using bootstrapping (Efron and Tibshirani, 1993; Lv et al., 2007; Chesnaye et al., 2021b). The T2-f-diag statistic, derived from the Hotelling-T2, assumes that frequency-based features are uncorrelated to estimate the test statistic (Chesnaye et al., 2023). Like the Q-Sample, the T2-f-diag sets the response threshold using bootstrapping, which is computationally intensive but highly accurate.

### 3.1. Simulation of neurophysiological responses in stationary and non-stationary noise

To evaluate detector performance, we conducted realistic simulations of cortical responses (Figure 2a) embedded in structured 1/*f* PSD noise, which approximates brain background activity (He et al., 2010). Simulations were performed under both stationary and non-stationary noise conditions, the latter incorporating eye-blink artefacts while maintaining all other parameters identical to the stationary condition. Parallel simulations without the target cortical signal (‘no-response’ data) were used to quantify detector sensitivity. First, we characterised the no-response data by examining the distribution of probability values (p-values) for each detector’s test statistic (the F ratio for parametric tests, Q- and T2-values for bootstrapped tests; see Methods). Ideally, these p-values should be uniformly distributed (Murdoch et al., 2008), indicating unbiased detection. Deviations from uniformity reflect bias; the larger the deviation, the greater the bias (Figure 2b). We also quantified false response detection rates as a function of the number of epochs, using a nominal sensitivity of 95% (Figure 2c).

In stationary noise simulations without a response, all detectors except Fmp-WAE and HT2-f-diag produced nearly unbiased estimates, as indicated by the flat p-value distributions (Figure 2b, stationary) and close agreement between empirical (black lines) and theoretical (orange lines) probability density functions (PDFs) of the F-distribution (insets in Figure 2b, stationary). In non-stationary noise, Fmp-WAE, Fmpi variants, and HT2 methods exhibited some bias (Figure 2b, non-stationary), which was also reflected in false response detection rates (Figure 2c). Under stationary noise (Figure 2c, left panel), Fmpi variants, HT2, HT2-f, Q-Sample, and T2-f-diag methods achieved nearly ideal false response detection rates of 5% (95% nominal sensitivity), both within and across all epochs (Figure 2c, left inset). The Fmp-WAE and HT2-f-diag detected fewer false responses than expected, indicating a bias towards higher sensitivity. In non-stationary noise, the Fmpi-WWE also showed a lower false response detection rate than the nominal value (Figure 2c, right panel). We then assessed true response detection rates by adding the simulated cortical responses to the noise. In stationary noise, final detection rates were: Fmpi variants (94.5-96.2%), bootstrapped Q-Sample (96.3%), Hotelling-T2 variants (87.2-95.8%), Fmp-WAE (71.8%), and bootstrapped T2-f-diag (96.0%) (Figure 2d, left). Under non-stationary noise, overall detection rates decreased (Figure 2d, right). The Fmpi-WWE outperformed other methods with a final detection rate of 54.9%, followed by the bootstrapped Q-Sample (48.4%). These detection rates were nearly double those of the remaining detectors, which ranged from 11.7% (Fmp-WAE) to 28.8% (HT2-f). Notably, the Fmpi-WWE showed higher detection rates despite a bias towards greater sensitivity in the absence of a response.

To further evaluate detector performance across different sensitivity values, we calculated several metrics. First, we compared nominal sensitivity (Figure 3a, solid black lines) to observed sensitivity (Figure 3a, coloured lines). Consistent with the initial results, observed sensitivity for all methods except Fmp-WAE and HT2-f-diag closely matched the nominal sensitivity under stationary noise (Figure 3a, left). The Fmp-WAE showed a better fit in non-stationary noise (Figure 3a, right panel).

Focusing on the clinically relevant sensitivity range (95-100%; Figure 3a, insets), the Fmp-WAE consistently showed a bias towards higher sensitivity in both stationary and non-stationary noise. The average bias (mean absolute difference between nominal and actual sensitivity) for stationary vs. non-stationary noise was: Fmp-WAE (2.57 vs. 1.33%), Fmpi-SA (-0.18 vs. 0.11%), Fmpi-WWE (0.05 vs. 1.63%), Fmpi-WAE (-0.28 vs. -0.12%), Q-Sample (-0.08 vs. 0.17%), HT2 (-0.19 vs. -0.98), HT2-f (-0.49 vs. -0.73%), HT2-f-diag (0.80 vs. 0.70 %), and T2-f-diag (-0.15 vs. 0.05 %), with the sign indicating a higher—positive—or lower—negative—average bias relative to the nominal value.

We then assessed performance as a function of the number of epochs (i.e., time) by generating sensitivity-specificity curves—ROCs—for the full sensitivity range (Figure 3b) and the clinically relevant range (Figure 3c). The highest performance over time—indicated by ROCs shifted towards the upper left corner—was observed for all Fmpi variants, bootstrapped Q-Sample, bootstrapped T2-f-diag, and frequency-based HT2 methods under stationary noise, and for Fmpi-WWE and bootstrapped Q-Sample under non-stationary noise. This pattern held across all sensitivity levels (Figure 3b) and within the clinically relevant range of the ROCs (Figure 3c), as shown by the AUC of the clinically relevant ROC (normalised by the total area within the relevant sensitivity region) over time (Figure 3d; see Methods).

We summarised overall performance by calculating the AAUC of the clinically relevant AUCs (Figure 3d) across all epochs (Figure 3e). AAUC values from 1000 simulations (treated as data from 50 participants) were analysed using a LME model, with AAUC as the dependent variable and detector type as the independent variable. The analysis revealed a significant effect of detector type [p < 0.001]. Post hoc comparisons confirmed that the Fmpi-SA, Fmpi-WAE, Q-Sample, and T2-f-diag significantly outperformed other detectors in stationary noise, whereas the Fmpi-WWE was superior under non-stationary noise (as indicated by non-overlapping error bars in Figure 3e).

To determine whether performance improvements were attributable to differences in observed sensitivity rather than intrinsic statistical properties of the detectors, thresholds for each detector were adjusted such that observed and nominal sensitivities were equal within the clinically relevant range. Following this equalisation the AUC remained largely unchanged under stationary noise, with improvements observed only for the Fmp-WAE and HT2-f-diag. This outcome was anticipated, as the Fmp-WAE and HT2-f-diag measure the same statistical quantities as the Fmpi-WAE and T2-f-diag, respectively. Under non-stationary noise, equalisation yielded results similar to the original analysis, with the notable exception of Fmpi-WWE, whose final AUC improved from 49.5 (unequalised) to 53.4% (equalised).

The generalisability of the Fmpi to other EP types was confirmed via simulations of ABRs (Sup. Figure 1) and ASSRs (Sup. Figure 2). The results indicated that Fmpi variants and Q-Sample approaches consistently yielded similar or superior performance relative to other detectors.

Having demonstrated the reliability of the Fmpi as a detector, its capacity for futility detection was systematically tested (see Methods). Specifically, we quantified the instances in which the estimated number of epochs to detect a cortical response, shown as a function of the actual number of epochs (Figure 4; each trace corresponds to an individual simulation), exceeded a predefined futility threshold of 200 epochs (Figure 4, horizontal line). In the presence of a cortical response under stationary noise (Figure 4, top-left panel), the trajectory of the estimated necessary epochs vs. the actual number of epochs typically crossed below the unitary diagonal—the detection boundary (Figure 4, diagonal black solid line)—prior to reaching the futility threshold, indicating that, in most simulations, the number of averaged epochs was sufficient to yield a significant response before futility was reached (Figure 4, green traces). Conversely, under non-stationary noise conditions (Figure 4, top-right panel), the futility threshold was surpassed at a substantially higher rate. That finding is consistent with the observed response detection rates (Figure 2d), wherein ≈ 94.5% of responses were detected at 200 epochs under stationary noise, compared to only ≈ 54.9% detected by Fmpi-WWE under non-stationary noise.

**Figure 4.**
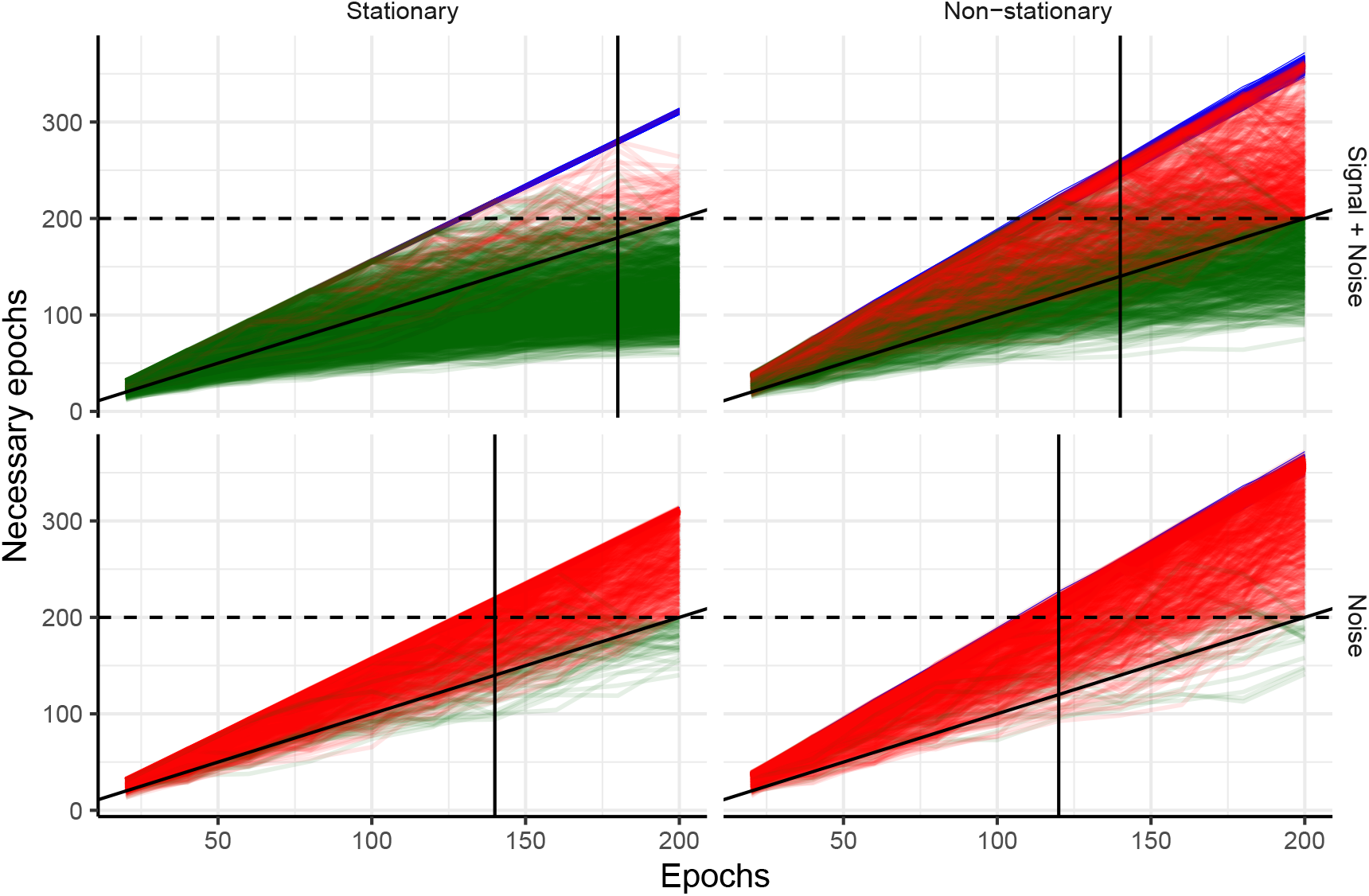
Fmpi enables reliable prediction of futility in simulated cortical recordings. The horizontal axes indicates the ongoing number of recorded epochs, whilst the vertical axes show the estimated number of epochs required to detect a significant cortical response. Results are based on 1000 simulations (each line corresponds to an individual simulation from Figure 2). Top panels show cortical simulations with Signal + Noise; bottom panels show, noise only data. Columns represent stationary and non-stationary noise. Green traces depict the trajectory of the estimated epochs required to detect a significant response, where detection occurred at the end of the simulation (at 200 epochs). Red traces indicate simulations in which a cortical response was not detected. Blue traces show the upper limit of the estimation for each simulated recording, determined by the absence of a neural response (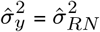, right side of Eq. 18). The diagonal black solid line marks the detection boundary: if the predicted number of epochs falls below the diagonal, the response has been detected; if it lies between the region determined by the noise (blue) and the diagonal, the response has not been detected. When the predicted number of epochs crosses the futility limit (horizontal black dashed lines), waiting for a response is considered futile. This estimation assumes that the overall recording conditions remain unchanged. Vertical black solid lines indicate the median number of epochs at which the futility threshold was exceeded. Traces were obtained using the Fmpi-WWE approach at a sensitivity of 95%.

To further assess the efficacy of the futility detector, we calculated the proportion of ‘expected’ futile measurements, defined as the ratio between the number of no detected responses at the end of the measurement period (at a nominal sensitivity of 95%) and the total number of measurements. For cortical responses in stationary noise, the expected futility was 5.5% whilst the observed futility rate—defined as the proportion of responses crossing the futility threshold at any time—was 7.8%, yielding a difference of 2.3% between predicted and observed futility. In non-stationary noise, the expected and observed futility rates were 45.1 and 53.0%, respectively. In the absence of a simulated cortical response, the expected and observed futility rates were 95.4 and 96.1% in stationary noise, and 97.9 and 99.0% in non-stationary noise, respectively. Absolute agreement between observed and predicted futility across all data, i.e., stationary and non-stationary noise with and without cortical responses, was 97% and was significant, as confirmed by Cohen’s Kappa coefficient (*κ*; Cohen, 1960) of 0.94 [p < 0.001].

Practically, these results imply that, when cortical responses are absent, half of the recordings would have terminated after 140 epochs (the median) in stationary noise and after 120 epochs in non-stationary noise, corresponding to 30% and 40% time savings, respectively, while maintaining error rates at the specified test sensitivity. Comparable results were obtained in simulations of ABRs and ASSRs (data not shown).

### 3.2. Detection of neurophysiological responses in human recordings

Having shown that the Fmpi and bootstrapped Q-Sample performed comparably, and frequently outperformed other detectors in simulated data, we subsequently evaluated their performance using EEG recordings from human participants. Specifically, EPs elicited by auditory stimuli from human participants across ten different studies (Table 1). EPs included ABRs, ASSRs, and cortical responses, with data collected from both adults and infants, including individuals with and without hearing loss, in a variety of clinical and research contexts. While the specific experimental conditions are detailed in Table 1, for the purposes of this analysis, only conditions in which EPs were expected were selected.

For time-domain analyses (i.e., ABRs and cortical responses), no-response datasets—required for estimating sensitivity, false response detection rates, and AAUCs—were generated by removing the EP from the original recordings and substituting the original event times with randomised events (see Methods). For frequency-domain analyses (i.e., ASSRs), the original event times were replaced with randomised ones and no-response analyses were carried out on neighbouring frequency bins adjacent to the target frequency, where no response was anticipated.

First, we analysed cortical EPs evoked by changes in ongoing stimuli (studies 1-3, ACC responses from adults) and by speech-like sounds (study 4; onset cortical responses from infants using hearing aids). The magnitude of the responses, which was influenced by stimulus characteristics, participants’ hearing status and age, as well as the total number of epochs, varied across studies (Figure 5a).

**Figure 5.**
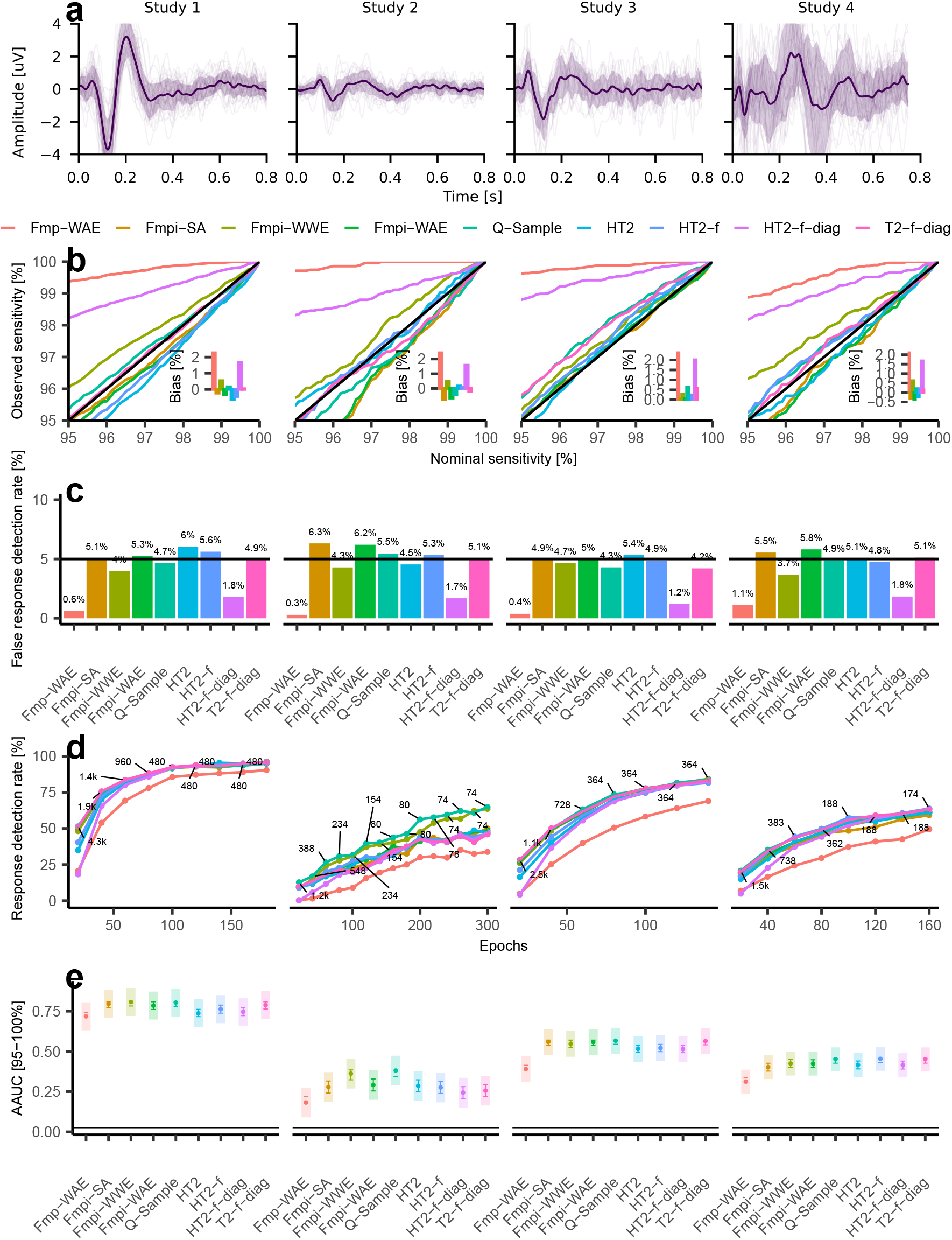
Fmpi enables robust detection of human cortical responses. **a** Example final averaged cortical EPs (bold lines) with individual data (thin lines) and standard deviation (shaded area) from different studies (each column represents a different study). **b** Nominal versus observed sensitivity for clinically relevant nominal sensitivity thresholds (95-100%). Traces represent data pooled across all epochs. The diagonal solid black line indicates the ideal unbiased expectation. Insets show the average bias across nominal sensitivities. A positive bias indicates that the obtained sensitivity was higher than the nominal value, whilst a negative bias indicates the opposite. **c** False response detection rates (derived from the same data) obtained for a nominal sensitivity of 95% pooled across all epochs. **d** Response detection rates for each study as a function of the number of epochs. The number of independent measurements contributing to each epoch is indicated by the numbers connected to the data points. **e** AAUC for the clinically relevant nominal sensitivities. Shaded areas represent 95% confidence intervals. The length of the error bars has been adjusted for visual post hoc comparisons (Holm FWER corrected). Non-overlapping segments indicate significant differences, whilst overlapping segments indicate non-significant differences. The horizontal solid black line indicates the chance level of the AAUC. The different methods are indicated by the colours across all panels.

Consistent with simulation results, the greatest sensitivity bias was observed for the Fmp-WAE and HT2-f-diag detectors (Figure 5b). In contrast, all other methods exhibited a small bias, averaging less than 1% (Figure 5b, insets). This trend was also reflected in the false response detection rates at a nominal sensitivity of 95% (Figure 5c). Notably, both the response detection rates at 95% nominal sensitivity (Figure 5d) and the AAUC (Figure 5e)—which summarises performance across epochs for all clinically relevant sensitivities—were highest for the Fmpi-WWE and Q-Sample methods.

This finding was corroborated by an LME model applied independently to each study, with AAUC as the dependent variable, *detector* as the fixed effect, and *subject* as a random effect. In all studies, the *detector* factor was significant [p < 0.001]. Post hoc comparisons, adjusted for FWER using the Holm method, indicated that the Fmpi-WWE and Q-Sample detectors consistently achieved the highest AAUC across all studies. No significant differences were observed between the Fmpi-WWE and Q-Sample methods in any study (as indicated by overlapping error bars in Figure 5e), and both methods performed significantly better than, or comparable to, the best of the remaining methods. The Fmp-WAE approach consistently exhibited the lowest performance, whilst the performance of the various HT2 methods and bootstrapped T2-f-diag varied by study.

In study 1, the Fmpi-WWE and Q-Sample outperformed the Fmp-WAE, HT2 and HT2-f-diag [p < 0.05]. In study 2, the Fmpi-WWE and Q-Sample outperformed all HT2 variants and bootstrapped T2-f-diag [p < 0.05]. In study 3, all Fmpi variants, bootstrapped Q-Sample, and T2-f-diag performed similarly [p > 0.05], although the Q-Sample method outperformed all HT2 variants. In study 4, only the Fmp-WAE detector demonstrated poor performance [p < 0.05]. Notably, results from study 2 paralleled findings from non-stationary noise simulations, demonstrating improved performance for the Fmpi-WWE and Q-Sample (cf. Figure 5d and Figure 2d).

These results confirm that the detection of cortical EPs—characterised by longer and fewer epochs—is most effectively achieved using the Fmpi-WWE, with performance equivalent to the more computationally intensive bootstrapped methods.

Subsequently, the detection methods were applied to ABRs (Figure 6a) obtained from both adults (studies 5 and 7) and infant participants (study 6). This dataset represented the largest and most computationally demanding cohort analysed, comprising ≈ 3000 independent measurements. The computational burden was primarily attributable to the bootstrapped methods (Q-Sample & T2-f-diag), which required substantial processing time; for instance, processing a single block of 4000 epochs necessitated over 50 seconds using six parallel processes, whereas parametric detectors (Fmp-WAE, Fmpi, and HT2 variants) completed the same task in a fraction of a second on a single processor.

**Figure 6.**
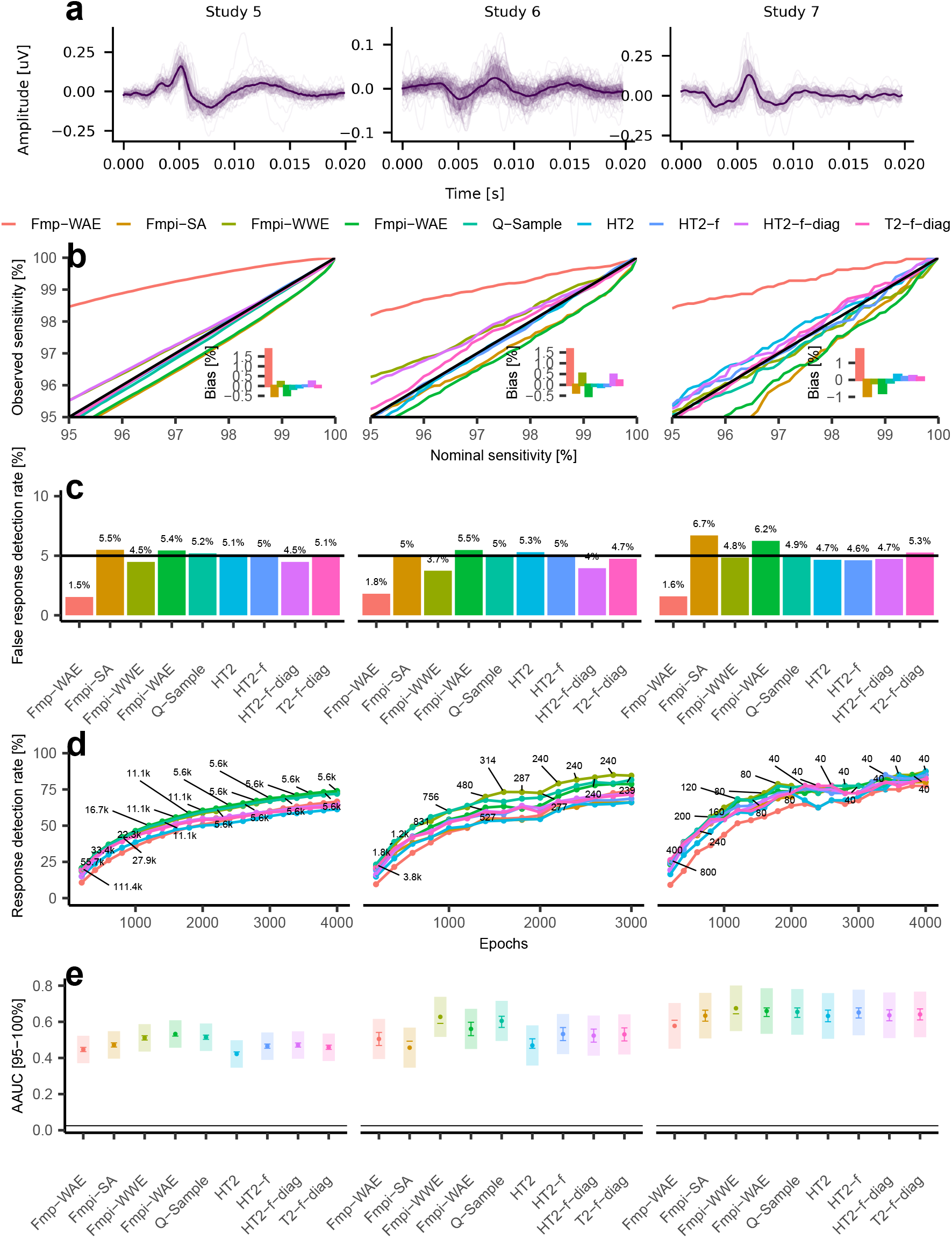
Fmpi enables robust detection of human brainstem responses. **a** Example averaged ABRs (bold lines) with individual data (thin lines) and standard deviation (shaded area) from different studies (each column represents to a different study). **b** Nominal versus observed sensitivity for the clinically relevant nominal sensitivities (95-100%). Traces represent data pooled across all epochs. The diagonal solid black line indicates the ideal unbiased expectation. Insets show the average bias across nominal sensitivities. A positive bias indicates that the obtained sensitivity was higher than the nominal value, whilst a negative bias indicates the opposite. **c** False response detection rates (derived from the same data) obtained for a nominal sensitivity of 95% pooled across all epochs. **d** Response detection rates for each study as a function of the number of epochs. The number of independent measurements contributing to each epoch is indicated by the numbers connected to the data points. **e** AAUC for the clinically relevant nominal sensitivities. Shaded areas correspond to 95% confidence intervals. The length of the error bars has been adjusted for visual post hoc comparisons (Holm FWER corrected). Non-overlapping segments indicate significant differences, whilst overlapping segments indicate non-significantly differences. The horizontal solid black line indicates the chance level of the AAUC. The different methods are indicated by the colours across all panels.

The observed bias (Figure 6b) and false response detection rates (Figure 6c) were analogous to those identified in the cortical response analyses. However, the HT2-f-diag approach exhibited improved performance (small degree of bias) compared to the Fmp-WAE method. The influence of the *detector* factor on AAUC was substantiated by an LME model [p < 0.001 across all three studies]. In accordance with simulation results, the Fmpi-WWE and Fmpi-WAE demonstrated superior performance among all Fmpi variants (Figure 6d&e). The performance of the Fmpi-WWE and Fmpi-WAE did not differ significantly from that of the Q-Sample method [p > 0.05 in all three studies, as indicated by overlapping error bars in Figure 6e]. Both Fmpi-WWE and Q-Sample methods significantly outperformed Fmpi-SA, Fmp-WAE, HT2, HT2-f, HT2-f-diag, and T2-f-diag [p < 0.05] in studies 5 and 6, whereas, in study 7, only the Fmp-WAE exhibited poor performance.

These results confirm that ABR detection is most effectively achieved using weighted averaging approaches (Fmpi-WAE and Fmpi-WWE).

Following the demonstration of the Fmpi’s robust performance with neurophysiological responses in the time domain, its efficacy in detecting ASSRs in the frequency domain was evaluated. Analyses encompassed ASSRs at high modulation rates (39 to 81 Hz; Figure 7), which are commonly employed in clinical practice, as well as at low modulation rates (≈ 7 Hz; Figure 8). The dataset included normal hearing (NH) adults (studies 8 and 9) and hearing-impaired (HI) infants using hearing aids (study 10). All detectors were provided with identical frequency sets: the modulation rate and the next two harmonics (studies 8 and 9), and the next eleven harmonics (study 10; see Methods).

**Figure 7.**
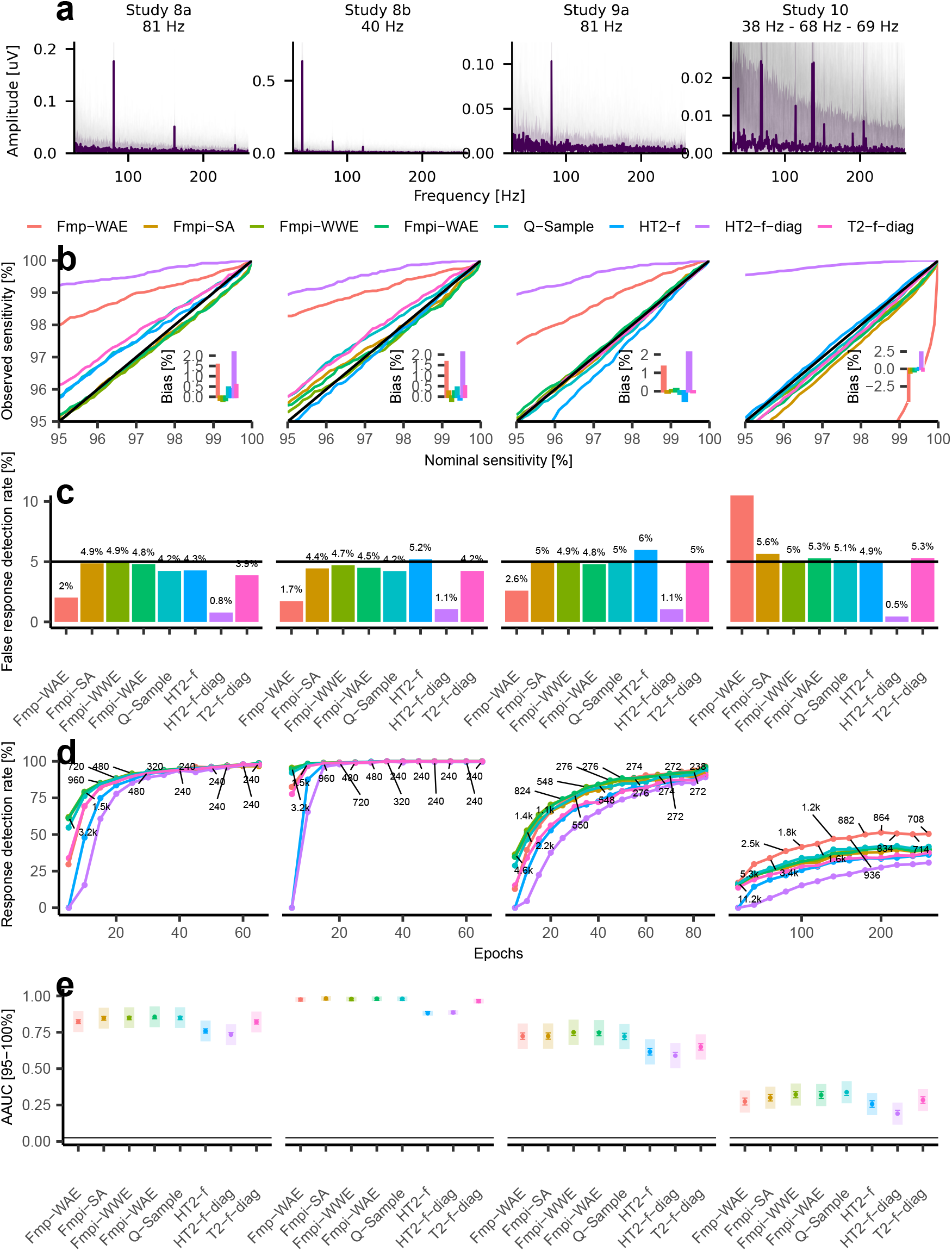
Fmpi enables robust detection of human steady-state responses at high modulation rates. **a** Example ASSRs from different studies (each column represents a different study). Responses were obtained for ASSR rates of ≈ 40 and 81 Hz (study 8), 81 Hz (study 9), and 38, 68 and 69 Hz, simultaneously (study 10). Three harmonics were included in studies 8 and 9, whilst 12 harmonics were included in study 10. **b** Nominal versus observed sensitivity for the clinically relevant nominal sensitivities (95-100%). Traces represent data pooled across all epochs. The diagonal solid black line indicates the ideal unbiased expectation. Insets show the average bias across nominal sensitivities. A positive bias indicates that the obtained sensitivity was higher than the nominal value, whilst a negative bias indicates the opposite. **c** False response detection rates (derived from the same data) obtained for a nominal sensitivity of 95% pooled across all epochs. **d** Response detection rates for each study as a function of the number of epochs. The number of independent measurements contributing to each epoch is indicated by the numbers connected to the data points. **e** AAUC for the clinically relevant nominal sensitivities. Shaded areas correspond to 95% confidence intervals. The length of the error bars has been adjusted for visual post hoc comparisons (Holm FWER corrected). Non-overlapping segments indicate significant differences, whilst overlapping segments indicate non-significantly differences. The horizontal solid black line indicates the chance level of the AAUC. The different methods are indicated by the colours across all panels.

**Figure 8.**
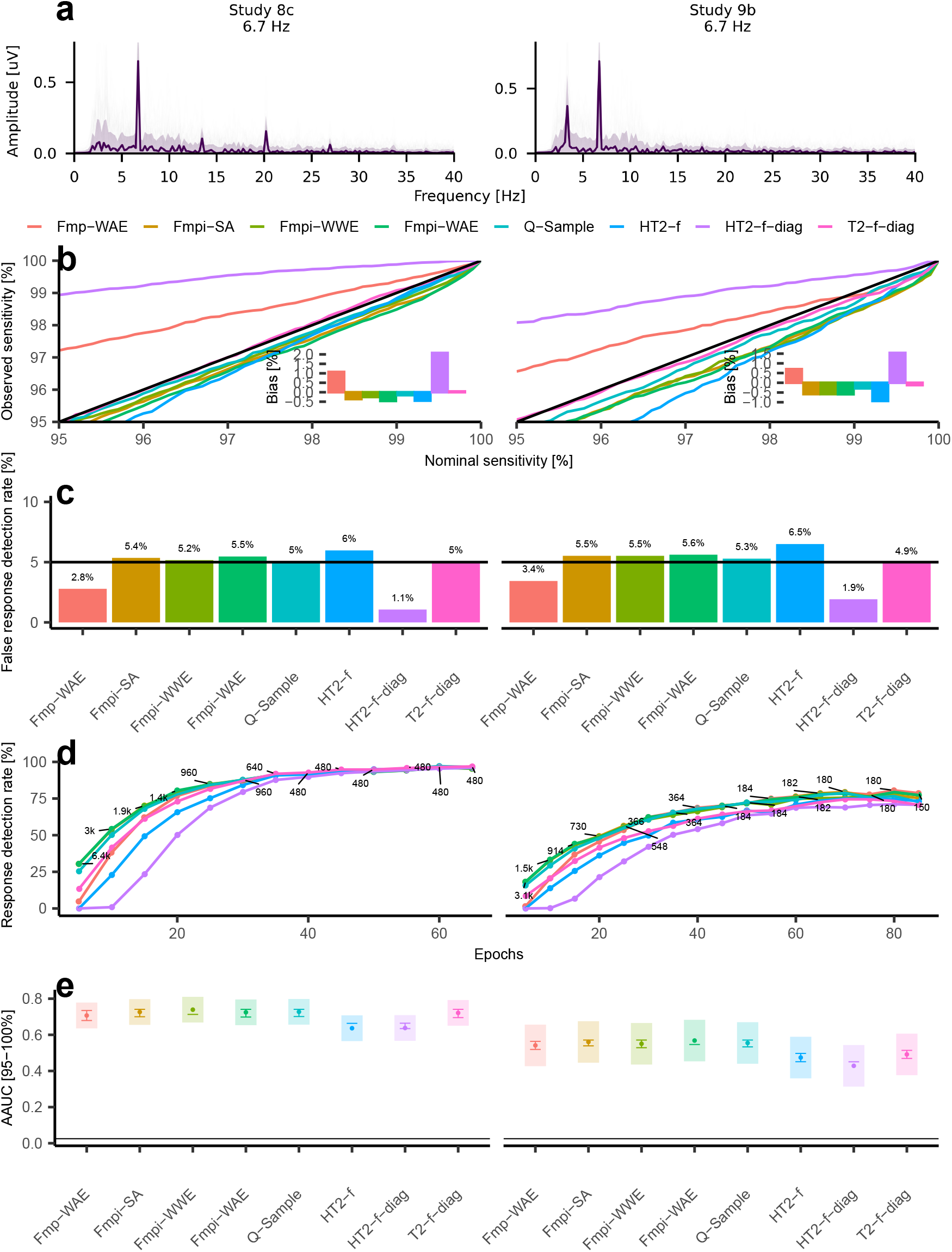
Fmpi enables robust detection of human steady-state responses at low modulation rates. **a** Example averaged ASSRs (bold lines) with individual data (thin lines) and standard deviation (shaded area) from different studies (each column represents a different study). Responses were obtained for ASSR rates of ≈ 7 Hz and the next two harmonics. **b** Nominal versus observed sensitivity for the clinically relevant nominal sensitivities (95-100%). Traces represent data pooled across all epochs. The diagonal solid black line indicates the ideal unbiased expectation. Insets show the average bias across nominal sensitivities. A positive bias indicates that the obtained sensitivity was higher than the nominal value, whilst a negative bias indicates the opposite. **c** False response detection rates (derived from the same data) obtained for a nominal sensitivity of 95% pooled across all epochs. **d** Response detection rates for each study as a function of the number of epochs. The number of independent measurements contributing to each epoch is indicated by the numbers connected to the data points. **e** AAUC for the clinically relevant nominal sensitivities. Shaded areas correspond to 95% confidence intervals. The length of the error bars has been adjusted for visual post hoc comparisons (Holm FWER corrected). Non-overlapping segments indicate significant differences, whilst overlapping segments indicate non-significantly differences. The horizontal solid black line indicates the chance level of the AAUC. The different methods are indicated by the colours across all panels.

Consistent with time-domain analyses, the greatest sensitivity bias was observed for the Fmp-WAE and HT2-f-diag detectors (Figure 7b&c and Figure 8b&c). This effect was particularly pronounced in study 10, where the Fmp-WAE—which assumes that each frequency contributes with equal DOF (see Methods)—resulted in reduced sensitivity, increased false response detection rates, and inflated detection rates (Figure 7b-d, study 10). Irrespective of modulation rate, the results corroborated simulation findings (Sup. Figure 2) with Fmpi variants and bootstrapped Q-Sample demonstrating superior performance (Figure 7d&e and Figure 8d&e).

LME modelling confirmed a significant effect of *detector* type on AAUC [p < 0.001 in all studies]. For high modulation rates ASSRs (Figure 7), post hoc comparisons revealed no significant differences among Fmpi-WWE, Fmpi-WAE, and Q-Sample (as indicated by overlapping error bars in Figure 7e). In all studies, the Fmpi-WAE, Fmpi-WWE, and bootstrapped Q-Sample outperformed HT2-f and HT2-f-diag [p < 0.05]. Additionally, in all but one study (study 8b), Q-Sample outperformed the bootstrapped T2-f-diag [p < 0.05], whilst Fmpi-WWE outperformed bootstrapped T2-f-diag in studies 8a and 9a [p < 0.05].

The results for low modulation rate ASSRs were analogous, with no significant differences between Fmpi variants and bootstrapped Q-Sample methods [p > 0.05; Figure 8a-e]. Both methods outperformed, or at least matched, the best performance of the remaining approaches. Notably, Fmpi maintained consistently high performance across a broad range of harmonics included in the analysis (1 to 12; Sup. Figure 3).

Collectively, these findings indicate that Fmpi is a robust general-purpose method, characterised by well-controlled sensitivity and consistently high performance as reflected by AAUC. Importantly, this approach imposes minimal computational demands on existing clinical systems while maintaining performance comparable to bootstrapped methods.

### 3.3. Testing time and futility detection in human recordings

To further assess the clinical utility of the Fmpi detector, overall testing time—defined in terms of the number of epochs—was quantified for each study and detection method. Testing times were estimated from the response detection rates derived for each study, calculated as a function of the number of epochs for nominal sensitivities ranging from 95% to 100% in increments of 0.01%. These response detection rate functions, which describe the proportion of detected responses as a function of accumulated epochs at a given sensitivity level, were used to estimate the number of responses detected at each epoch from an initial pool of 10000 measurements. Once a response was detected, the corresponding measurement was removed from the pool, and no further testing was assumed. For each nominal sensitivity, testing times for all detectors were compared relative to the fastest detector, and the difference was normalised and expressed as a percentage of the shortest testing time. Positive values thus indicate additional testing time incurred relative to the fastest detector at a given sensitivity level. The mean additional testing time across all sensitivity levels was subsequently calculated to provide an overall estimate of average performance. To ensure fairness and comparability across detectors, reductions in testing time attributable to futility detection—which is only available for the Fmpi detector—were excluded from this analysis.

Consistent with their superior response detection rates, both Fmpi and bootstrapped Q-Sample detectors exhibited the shortest average testing times across all studies (Figure 9a; Table 2). For cortical responses, the average additional testing times were as follows: Fmpi-SA (8.8%), Fmpi-WWE (4.9%), Fmpi-WAE (6.8%), Q-Sample (0.3%), and T2-f-diag (7.1%), all of which were substantially shorter than those for Fmp-WAE (63.0%), HT2 (20.9%), HT2-f (13.8%), and HT2-f-diag (33.4%). For ABRs, Fmpi-WAE (3.3%), Fmpi-WWE (2.6%) and Q-Sample (1.1%) outperformed Fmp-WAE (44.2%; the clinical standard), Fmpi-SA (7.5%), HT2 (23.2%), HT2-f (8.8%), HT2-f-diag (11.2%), and T2-f-diag (7.2%). In the context of ASSRs, Fmpi-SA (5.8%), Fmpi-WWE (3.7%), Fmpi-WAE (4.6%), and Q-Sample (10.3%) again demonstrated superior efficiency compared to Fmp-WAE (27.7%), HT2-f (73.5%), HT2-f-diag (114.5%), and T2-f-diag (34.2%). These findings indicate that the Fmpi detector can substantially reduce testing time, achieving efficiency comparable to the computationally intensive bootstrapped methods, without compromising sensitivity or performance.

**Table 2.** Additional detection times. Columns, from left to right: study ID; EP type; and the various detection methods. The additional detection time (in %) was estimated as the ratio of the testing time for each detector to the shortest detection time, calculated for nominal sensitivities between 95 and 100% (in increments of 0.01%, excluding 100%). The standard deviation is shown in parentheses (in %).

| Study | EP | Fmp-WAE | Fmpi-SA | Fmpi-WWE | Fmpi-WAE | Q-Sample | HT2 | HT2-f | HT2-f-diag | T2-f-diag |
| --- | --- | --- | --- | --- | --- | --- | --- | --- | --- | --- |
| 1 | Cortical | 50.2 ( 6.9) | 0.7 ( 0.7) | 2.7 ( 0.8) | 0.4 ( 0.7) | 0.5 ( 0.5) | 19.4 ( 3.3) | 12.9 ( 3.0) | 33.2 ( 0.7) | 0.4 ( 0.8) |
| 2 | Cortical | 100.1 ( 7.1) | 27 ( 2.0) | 8.7 ( 2.0) | 22.4 ( 1.5) | 0 ( 0.0) | 30.9 ( 2.4) | 23.4 ( 2.3) | 48.9 ( 4.2) | 25.1 ( 1.1) |
| 3 | Cortical | 57.5 ( 3.3) | 0.7 ( 0.8) | 2.7 ( 0.6) | 0.5 ( 0.8) | 0.8 ( 0.9) | 20.2 ( 3.1) | 12.6 ( 2.4) | 28.2 ( 1.9) | 1.4 ( 1.3) |
| 4 | Cortical | 44.3 ( 4.3) | 6.6 ( 1.4) | 5.6 ( 2.2) | 3.8 ( 1.5) | 0 ( 0.2) | 13 ( 1.5) | 6.3 ( 1.8) | 23.5 ( 4.0) | 1.3 ( 0.6) |
| 5 | ABR | 35.5 ( 3.5) | 2.8 ( 0.6) | 5.3 ( 0.4) | 0 ( 0.4) | 2.5 ( 1.1) | 21 ( 2.1) | 6.9 ( 1.2) | 9.6 ( 0.4) | 6.4 ( 1.3) |
| 6 | ABR | 44.1 ( 4.2) | 16 ( 1.0) | 1.2 ( 1.0) | 9.3 ( 0.8) | 0.1 ( 0.4) | 26.5 ( 2.3) | 12.8 ( 1.3) | 14 ( 1.2) | 11.1 ( 1.4) |
| 7 | ABR | 53.1 ( 6.3) | 3.8 ( 0.9) | 1.4 ( 0.9) | 0.7 ( 1.0) | 0.8 ( 1.0) | 22 ( 1.8) | 6.5 ( 1.9) | 10 ( 1.0) | 4.3 ( 1.7) |
| 8a | ASSR | 42.5 ( 6.7) | 1.7 ( 0.3) | 0 ( 0.0) | 1.2 ( 0.2) | 10.4 ( 2.7) | NA | 86.5 ( 4.4) | 126 ( 2.3) | 37.2 ( 5.8) |
| 9a | ASSR | 34.2 ( 1.9) | 5.7 ( 0.7) | 0 ( 0.0) | 2.7 ( 0.3) | 14 ( 2.1) | NA | 71.9 ( 3.3) | 118.5 ( 1.6) | 43.4 ( 3.3) |
| 8b | ASSR | 29.1 (17.9) | 0.8 ( 0.2) | 0 ( 0.0) | 0.8 ( 0.2) | 6.3 ( 3.4) | NA | 113.8 (12.5) | 146.4 (17.4) | 33.3 (14.3) |
| 10 | ASSR | 0 ( 0.0) | 25.4 ( 2.1) | 21.5 ( 1.8) | 22.9 ( 1.9) | 17.2 ( 1.4) | NA | 62.8 ( 1.9) | 111.2 ( 9.4) | 36.1 ( 2.8) |
| 8c | ASSR | 34.1 ( 1.0) | 0.4 ( 0.4) | 0.2 ( 0.4) | 0 ( 0.3) | 7.7 ( 1.5) | NA | 56.8 ( 1.0) | 97 ( 3.3) | 26.4 ( 2.3) |
| 9b | ASSR | 26.3 ( 1.1) | 0.6 ( 0.7) | 0.4 ( 0.6) | 0.2 ( 0.6) | 6.5 ( 1.4) | NA | 49.3 ( 2.2) | 87.7 ( 3.9) | 29 ( 2.4) |

**Figure 9.**
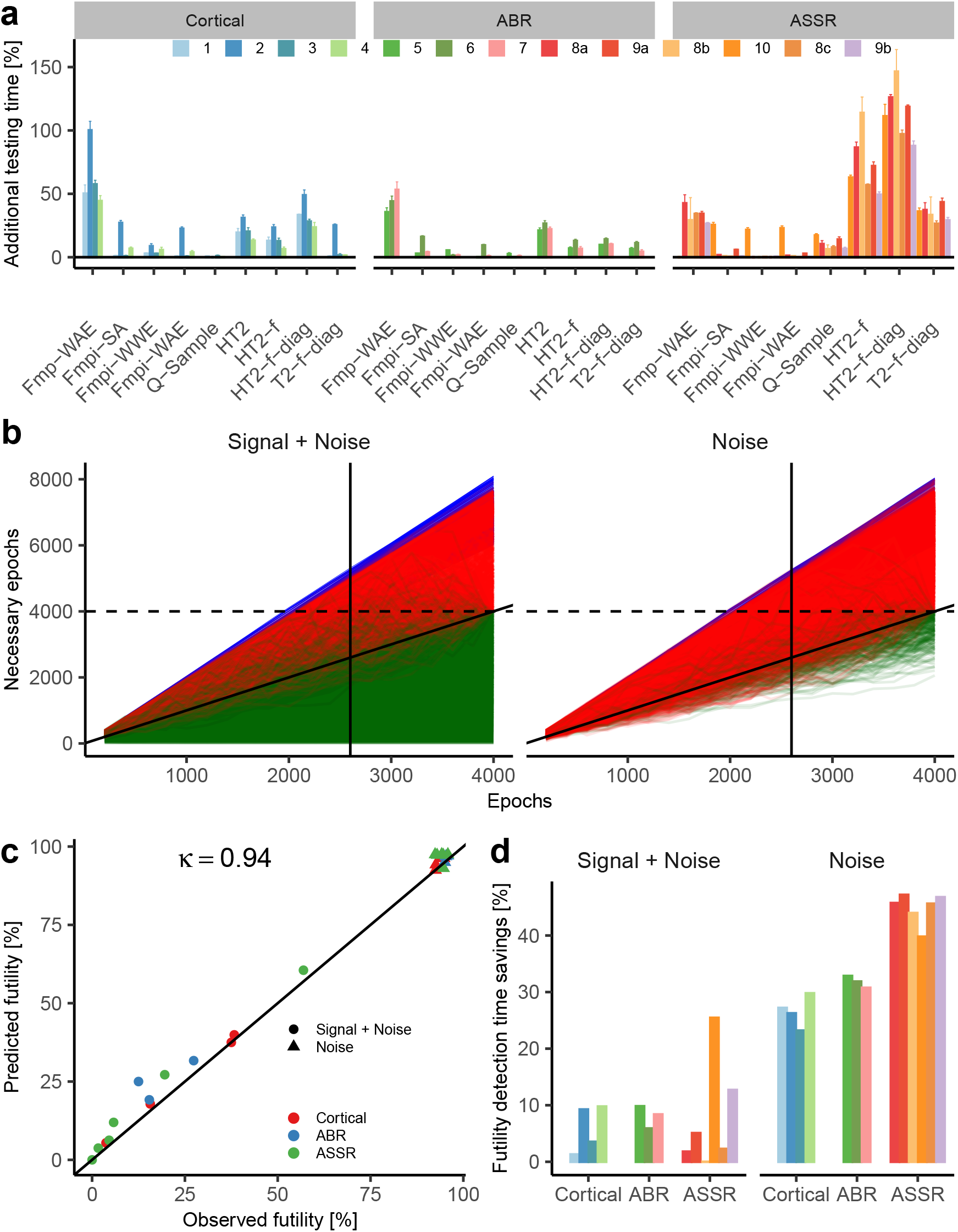
Fmpi significantly reduces testing time for neurophysiological response detection. **a** Normalised testing times for each detection method grouped by EP type (Cortical, ABR, and ASSR). Normalised times are averaged across all clinically relevant nominal sensitivities (95-100%). Study identifiers are indicated by colour. **b** Futility curves for ABRs from study 5 (n = 5570 traces). The left panel shows results for original data (Signal + Noise), whilst the right panel present results for no-response data derived from the same dataset (Noise). Green traces depict the trajectory of the ongoing (horizontal axis) versus the predicted (vertical axis) number of epochs for each measurement in which a response was detected at the end of the recording (4000 epochs). Red traces indicate measurements where the futility threshold was exceeded (horizontal dashed line). Blue traces show the upper limit of the estimation (determined by the noise; right side of Eq. 18) for each recording. The diagonal solid black line denotes the detection boundary. A trace falling below the diagonal indicates EP detection. Vertical black solid lines indicate the median number of epochs at which futility was detected. **c** Relationship between observed (horizontal axis) and predicted (vertical axis) futility rates across all studies. **d** Futility-based time savings across all studies, shown for both the presence (Signal + Noise) and absence (Noise) of an EP.

In addition to improved response detection time, the Fmpi offers further time savings through futility detection. This aspect was also investigated from EP recordings. Employing the same methodology as in the simulations, expected and predicted futility rates were compared based on a predefined futility threshold, defined as the total number of epochs available for each recording. Analyses were conducted for both the original data (Signal + Noise) and the no-response data derived from the same datasets (Noise). The futility curves derived from human data (Figure 9b; from study 5) closely paralleled those obtained in simulations (Figure 4).

The concordance between expected and predicted futility was highly significant (Figure 9c). Absolute agreement percentages (Table 3), reflecting the alignment between final detection and futility detection for each measurement, ranged from 87.5% to 100% across individual studies, with an overall agreement of 97.1% when aggregated across all studies. These findings were further supported by Cohen’s Kappa coefficient analyses, with *κ* values spanning 0.87 to 0.97 across studies [p < 0.001 for all cases], and a pooled *κ* value of 0.94 [p < 0.001]. Notably, *κ* values exceeding >0.80 are generally interpreted as ‘almost perfect’ agreement (Landis and Koch, 1977).

**Table 3.**
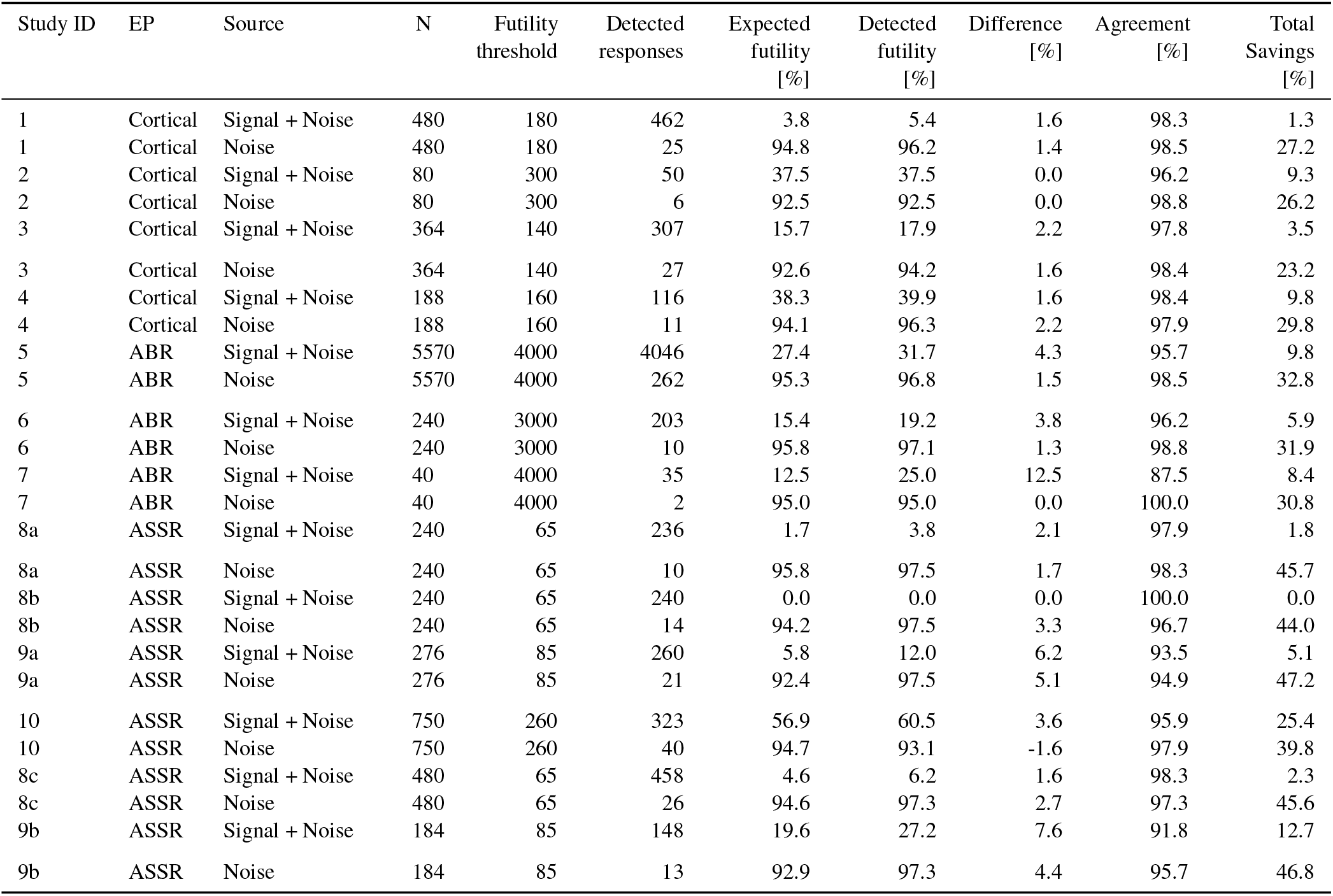
Futility detection across studies. Columns, from left to right: study ID; EP type; type of data analysed (original recording: Signal + Noise; or derived noise data: Noise); total number of measurements analysed (N); futility threshold (in epochs); number of detected EPs; expected futility (%) based on the total number of detected responses; detected futility (%); difference between expected and detected futility (%); agreement (%) between expected and observed futillity based on individual measurements; and total savings (as a percentage of epochs) based on measurements being terminated when futility was triggered (%). Analyses were conducted at a nominal sensitivity of 95% employing the Fmpi-WWE detector.

The proportion of time saved—defined as the ratio between the epoch at which futility was detected and the predefined futility threshold—varied across studies and datasets (Figure 9d). For the original recordings, time savings were inversely related to response detection rates, as expected. For instance, time savings of approximately 25% were observed for ASSRs in study 10 which exhibited a response detection rate of 43%, whereas savings were reduced to 13% for ASSRs in study 9b (80% detection rate) and to 9.8% for ABRs in study 5 (73% detection rate). In contrast, no time savings were observed for the 40 Hz ASSRs in study 8b, which demonstrated a response detection rate of 100%.

When the analysis was applied to no-response data, reflecting the maximal potential benefit of futility detection, time savings ranged from 23% for cortical recordings to 47% for ASSR data.

The results demonstrate that Fmpi significantly shortens the time required to detect the presence of neurophysiological responses or to determine when a response is unlikely to emerge, particularly in scenarios where responses must be obtained within a predefined time frame or the RN must fall below some clinically established value. Moreover, the detector is computationally efficient and compatible with existing clinical devices, offering the potential for meaningful improvements in clinical practice.

## 4. Discussion

In the present study, we introduced and validated a novel general framework for statistical inference under coloured noise in neurophysiological signals—the Fmpi—which explicitly accounts for the spectral and temporal structure of the background noise. By leveraging these properties, the method enables analytical estimation of the effective DOF (*v*_*y*_ and *v*_*rn*_) governing the F-distribution used for response detection. Across extensive simulations and human datasets, the Fmpi achieved sensitivity and specificity comparable to computationally intensive bootstrap-based approaches, while consistently outperforming existing parametric methods. These gains were observed in both time- and frequency-domain analyses, supporting the general applicability of the proposed approach.

Beyond statistical performance, the Fmpi substantially reduced testing time by enabling both earlier detection of responses and principled termination of recordings when responses are unlikely to emerge. Under the present analysis settings, reductions in detection time ranged from 6% to 111% across brainstem, midbrain, and cortical AEPs relative to other parametric methods, and were comparable to those obtained with the bootstrapped Q-Sample approach. Clinically, the method reduced ABR detection time by 42% relative to the current standard (Fmp-WAE).

Additional gains (23% to 47%), were achieved through futility detection, highlighting the potential of the framework to improve efficiency in clinical and experimental settings.

### 4.1. Individualised inference under structured noise

A central contribution of this work is the shift from fixed or empirically tuned parameters toward data-driven, individualised inference. Conventional parametric approaches such as Fmp-WAE rely on predefined DOF values that depend on acquisition parameters and noise characteristics, including sampling rate, filtering, and artefact contamination (see Sup. Figure 4a&b). Similarly, multivariate approaches such as Hotelling’s *T* ^2^ implicitly handle the noise structure captured by the covariance matrix of the features included in the analysis. In both cases, population-based parameter tuning is required to match specific recording conditions.

These adaptations are necessary because serial correlations within epochs—and thus the effective DOF—are strongly governed by the spectral properties of the underlying noise. However, such dependencies are rarely modelled explicitly. Instead, they are absorbed indirectly through parameter choices, limiting generalisability across paradigms and populations.

The Fmpi addresses this limitation by incorporating all available samples as features and directly estimating the effective DOF from the data. Crucially, this estimation accounts for both the spectral colour (e.g., 1/*f* structure) and non-stationary temporal dynamics of the noise. This enables the method to operate robustly across a wide range of acquisition settings without requiring retuning, while achieving performance comparable to fully data-driven bootstrap approaches.

Importantly, the estimated DOF values align closely with those reported in prior empirical studies. For example, analytical predictions derived from the Fmpi reproduce classical estimates reported by Elberling and Don (1984) under matched conditions, including both white and pink noise regimes. Specifically, Elberling and Don (1984) reported an empirical DOF *v*_1_ of 160 for a 10 ms time window, in stationary white noise high-pass filtered at 100 Hz, sampled at 16 kHz, and averaged across 250 epochs. Applying the Fmpi’s analytical solution for white noise under the same conditions yielded an identical value (see Sup. 1.13 and Sup. Figure 4b, green marker for *p =* 0). Similarly, under structured pink noise bandpass filtered between 200 and 1000 Hz, the same study reported an empirical value *v*_1_ of 15.0. The Fmpi’s analytical solution using ideal filters produced a *v*_1_ of 13.0 (Sup. Figure 4b, green marker for *p =* 1), and 15.0 when using steep, non-ideal filters (not shown).

Further validation is provided by the seminal work by Elberling and Don (1984), who estimated fixed DOF *v*_1_ values by measuring multiple realisations from non-stimulus recordings in adult human participants, yielding a range of 8 to 22 *v*_1_ DOF. These were subsequently fitted to a Gaussian distribution, with a fixed DOF *v*_1_ of 5 corresponding to the 5% percentile threshold. Employing the same approach, the 5th percentile of *v*_1_ estimated from adult ABR recordings (studies 5 and 7) was 6.07, ranging from 3.6 to 36.8 DOF.

In a recent publication, Lightfoot et al. (2023) estimated the 97.5% and 99% F-value percentiles as 2.2 and 2.8, respectively, from no-response data from infants (33-1500 Hz bandpass filtered; 10 ms time window). The estimates from infants’ data analysed here (study 6) were 2.1 and 2.5, respectively, whilst the 5th percentile of *v*_1_ was 6.3, ranging between 3.8 and 22.8 DOF.

The wide range of *v*_1_ values estimated by the Fmpi is comparable to those shown by Elberling and Don (1984) and Lightfoot et al. (2023), and underlines the importance of using individualised rather than fixed DOF.

A related perspective emerges when considering feature selection with the time-domain Hotelling-*T* ^2^ approach. Reported optimal feature counts—specifically, voltage mean intervals—for ABRs (30-1500 Hz bandpass filtered; 15 ms time window) in simulated data range from 16 (corresponding to an interval of 938 µs) to 40 (375 µs intervals) (Chesnaye et al., 2018; McKearney et al., 2022). Analytical calculations of *v*_1_, interpreted here as the effective number of independent samples within the same bandwidth and analysis window, yielded values between 14.1 (1 ms, pink noise) and 44 (340 µs, white noise).

In a previous study, cortical responses analysed with the time-domain Hotelling-*T* ^2^, employed voltage mean intervals of 50 ms (resulting in nine features for data bandpass filtered between 0.1 and 30 Hz, using a 450 ms analysis window; Golding et al., 2009), whilst recent studies have employed intervals ranging between 33 to 50 ms (Bardy et al., 2020; Chesnaye et al., 2021a; Chesnaye et al., 2023; Sohier et al., 2021; Ching et al., 2023). Analytical solutions for *v*_1_ using filter parameters and analysis windows similar to those described by Golding et al. (2009) produced time intervals ranging from 153 ms (2.93 DOF, pink noise) to 17 ms (16.72 DOF, white noise).

The 5th percentile of DOF *v*_1_ estimated from adult data in studies 1 to 3 was 10.8 (corresponding to 42.0 ms interval), which falls within the range reported in those investigations.

Based on these results, empirical feature optimisation can be interpreted as an indirect strategy for reducing covariance between samples. The Fmpi makes this relationship explicit by analytically linking DOF to spectral properties, thereby unifying parametric and multivariate perspectives on neurophysiological response detection.

### 4.2. Methodological considerations and parameter dependencies

To ensure meaningful comparison across detectors, analysis parameters were harmonised with those used in previous studies (Chesnaye et al., 2018; Chesnaye et al., 2023), without attempting dataset-specific optimisation. Additional analyses matching signal energy across time- and frequency-domain methods confirmed that the overall ranking of methods remained unchanged, with the Fmpi and Q-sample approaches consistently demonstrating superior performance. Frequency-domain HT2 methods exhibited a stronger sensitivity bias, though the general pattern of results was preserved.

A critical consideration in time-domain analyses concerns the interaction between analysis window duration and high-pass filtering. As previously noted (Elberling and Don, 1984; Don and Elberling, 1994), mismatches between the spectral content of the averaged response and the RN can bias the F-statistic. Specifically, when the high-pass filter cutoff is lower than the effective cutoff imposed by demeaning within the analysis window, the RN variance includes low-frequency components that are absent from the averaged signal. This imbalance leads to reduced detection rates, particularly in the presence of strong low-frequency noise components. Previous studies employing the Fmp-WAE method for cortical and ABR analyses have often overlooked this relationship (Chesnaye et al., 2018; Chesnaye et al., 2021a; McKearney et al., 2022; Lightfoot et al., 2023), potentially introducing biases. This issue is not specific to conventional methods but also applies to the Fmpi. It can be mitigated either by selecting filter cutoffs consistent with the analysis window (e.g., *f*_*HP*_ ≥ 1/*T*_*w*_) or by ensuring that both signal and noise variances are estimated over comparable temporal windows. These considerations highlight the importance of aligning preprocessing and statistical inference, and may warrant more explicit treatment in future methodological guidelines and clinical standards (e.g. those of the British Society of Audiology). Finally, in this study we intentionally refrained from applying epoch-rejection procedures to assess method robustness under non-stationary noise. Whilst rejection improved absolute performance—particularly for cortical responses—it did not alter the relative ranking of methods. This suggests that the advantages of the Fmpi are not contingent on aggressive artefact control, although such preprocessing may further enhance overall performance.

### 4.3. Limitations and further directions

Several limitations should be considered. First, as with all sequential testing procedures, appropriate correction for multiple comparisons is required to maintain control over false positive rates (Albers, 2019). Although such corrections were not applied in the present analyses, we expect the relative performance of the Fmpi to remain largely unaffected given its robustness across sensitivity regimes.

Second, the proposed futility detection framework relies on predefined measurement limits derived from normative data or, alternatively, from normative RN values. Whilst effective in practice, futility decisions remain probabilistic and should be interpreted in the context of recording quality. In cases of excessive noise or suboptimal acquisition conditions, retesting may be necessary to ensure reliable conclusions.

Third, the current formulation assumes independence of noise across epochs, an assumption typically enforced through high-pass filtering. However, this assumption may be violated in scenarios involving spatial filtering (e.g., denoising source separation, de Cheveigné and Simon, 2008) or multichannel integration, where dependencies can arise across both temporal and spatial dimensions. Extending the framework to explicitly model such dependencies represents an important direction for future work.

Finally, further optimisation of analysis parameters—including filter bandwidths and analysis windows—may yield additional performance gains. Systematic evaluation across a broader range of paradigms and modalities (e.g, fNIRS) will be important to fully characterise the generalisability of the approach.

## Data Availability

The raw electroencephalography datasets analysed in this study originate from multiple independent studies and are subject to ethical, institutional, and consent-related restrictions. As a result, the raw data are not publicly available. Access requests should be directed to the investigators responsible for each dataset, as listed in Table 1, and will be assessed in accordance with the relevant data-sharing agreements. Derived data supporting the findings are available from the corresponding author upon reasonable request.

## Code availability

All methods described in this study are implemented in pEEGy, an open-source Python package developed for the analysis and simulation of electroencephalography data and evoked responses. The pEEGy source code is publicly available and maintained under an open-source licence. The version used for the analyses reported here is archived with a persistent digital object identifier (10.5281/zenodo.19512653), enabling reproducibility of the results. Documentation and usage examples are provided with the software.

## ACKNOWLEDGMENTS

This study received no funding. The authors thank Linsey Van Yper, Lisbeth Birkelund Simonsen, Mats Rekswinkel, Sinnet Greve Bjerge Kristensen, Guy Lightfoot, Macarena Bowen, and Anisa Visram for allowing us to analyse their data. We also thank James Harte and Bue Kristensen for their support and inspiring discussions about this study.

## Author contributions

JU conceptualized and developed the novel Fmpi method. JU, MC, DS, and SL defined and conceptualised comparisons with other detection methods. JU analysed both simulations and neurophysiological data and wrote the manuscript with input from all authors.

## Supplementary information

For the next developments we will consider the following definitions of mean, *µ*, and variance, *σ*^2^, for a random variable *x*:

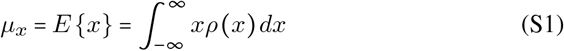

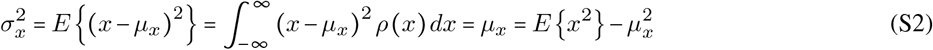

where *ρ*(*x*) is the probability density function of the variable x.

Consider a *χ*^2^ random variable with *n* DOF

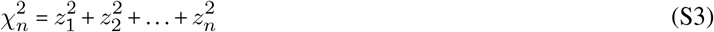

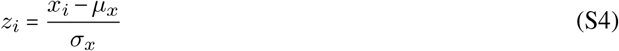

where *x*_*i*_ is random normally distributed variable and *z*_*i*_ is a standardised random variable with *µ*_*z*_ *=* 0 and 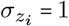. The following properties for *χ*^2^ random variables can be demonstrated.

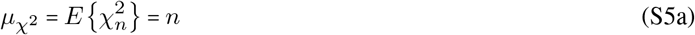

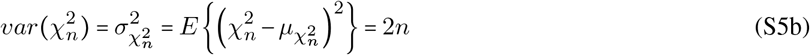

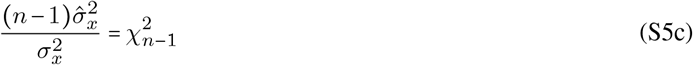

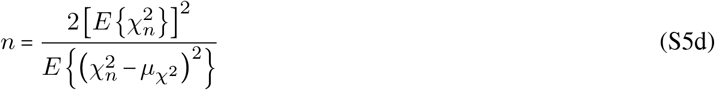

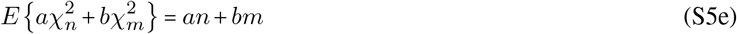

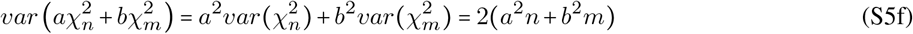

### Sup. 1.4. F-statistic

By definition, the F-statistic corresponds to the ratio between the variance of two independent random variables. Specifically for an EP, the F-statistic corresponds to the ratio between the variance of the averaged signal, across many epochs within a recording 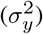—which contains both the signal of interest (*x*) and residual noise (*RN*)—and variance of the estimated residual noise 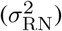.

If the signal of interest and the background noise are independent processes, the F-ratio (this is derived later) can be expressed as

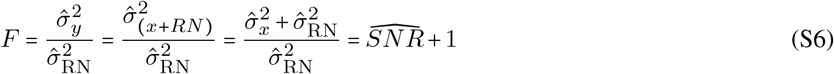

Note that F is the ratio of two variances, thus it can be parametrised by a Fisher–Snedecor distribution as *F* (*x, v*_*y*_, *v*_RN_) where *v*_*y*_ and *v*_RN_ are the DOF of the respective variances in Eq. S6.

The estimation of the residual noise variance 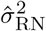 epends on the type of average utilised.

### Sup. 1.5. Response average

Let’s consider the following time series

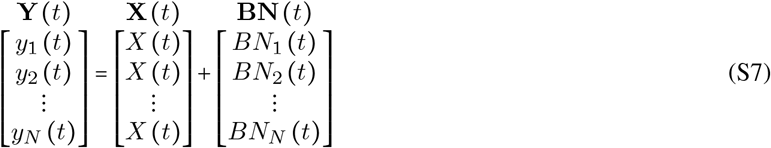

where *y*_*i*_ (*t*) is the *i*-th recorded epoch of a total of *N*, *X* (*t*) is the *true* response of interest, and *BN*_*i*_ (*t*) is the underlying background noise present on each epoch. For simplicity, we will assume that both *X* (*t*) and *BN*_*i*_ (*t*) have zero mean.

The average response estimate can be expressed as

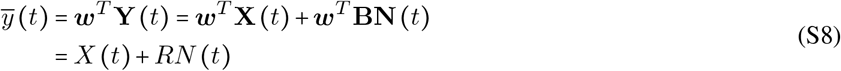

where *RN =* ***w***^*T*^ **BN** *t* corresponds to the post-averaged RN.

Using standard averaging, the weight vector ***w*** is given by

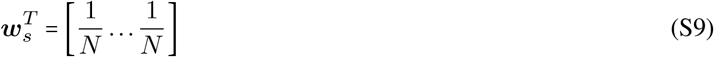

whereas for weighted averaging based on the inverse of the variance (Elberling and Don, 1984; Don et al., 1984; Don and Elberling, 1994), ***w*** is given by

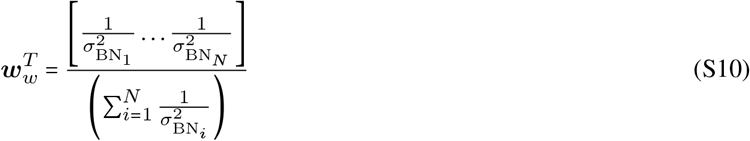

where 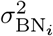 corresponds to the variance within each epoch.

### Sup. 1.6. SNR and F-value

The SNR of the measurement can be written as

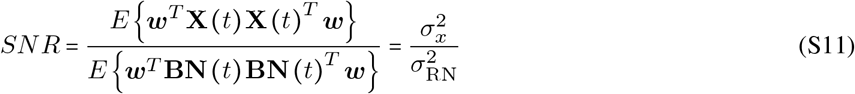

As shown in Eq. S11, the SNR corresponds to the variance of the target neural signal (*σ*^2^) and the variance of the RN 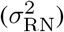.

The variance of the averaged recording 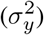—which includes both the target signal and the noise—can be written as

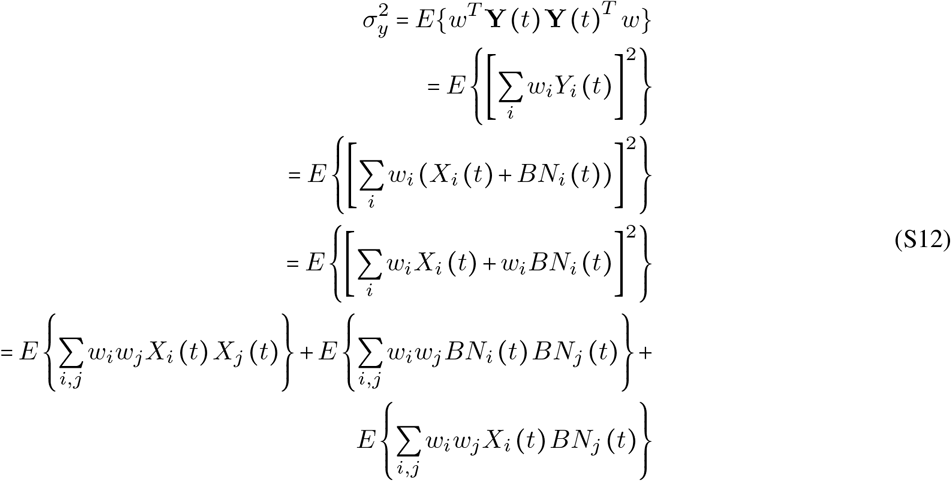

Under the assumption that the signal and the noise are independent processes (*E* {*X*_*i*_ (*t*)*BN*_*j*_ (*t*)} = 0), and that the noises at each epoch are also independent (*E* {*BN*_*i*_ (*t*)*BN*_*j*_ (*t*)}_*i*≠*j*_ = 0) this can be simplified as

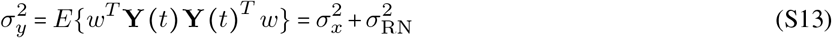

Therefore, Eq. S11 can be written as

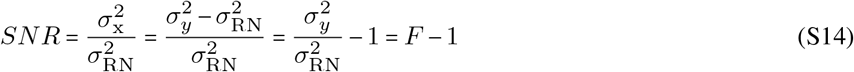

### Sup. 1.7. Estimation of the background noise variance across epochs

To compute the F-ratio we must find a reliable estimate of 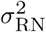, which in turn requires a reliable estimation of the underlying potentially non-stationary background noise variance 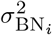. This can be achieved by means of two different methods.

The first method estimates 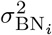 by tracking the variance of a single or multiple fixed time-points *across epochs* (Elberling and Don, 1984; Elberling and Wahlgreen, 1985; Don and Elberling, 1994; Silva, 2009). In this case, if the noise is a locally stationary ergodic process, the variance of such a local noise source can be calculated as

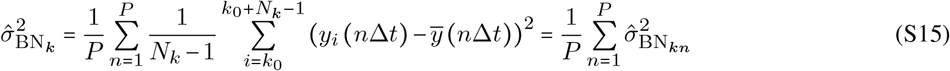

where *P* is the number of tracked points used to estimate the background noise, *N*_*k*_ is the number of epochs used to estimate the variance of each block, *k*_0_ is the starting epoch of the block, and Δ*t* is the time interval between the tracked samples.

This method assumes that the noise is locally stationary within each block of *N*_*k*_ epochs, that is, 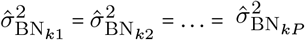.

### Sup. 1.8. Estimation of the background noise variance within epoch

This second method estimates the underlying background noise variance 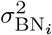 from the variance *within each epoch* as shown in Eq. S16.

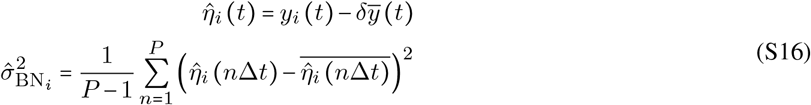

Here, *δ* is 0 or 1, pending on the assumption that the variance of the *true* target response 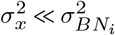, or not. In many real applications 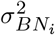 will 20 dB or more above 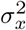. If that is the case, then the assumption will be true, and *δ* will be zero.

### Sup. 1.9. Estimation of the residual noise variance

Regardless of the chosen method to estimate the background noise (Eq. S15 or Eq. S16), the variance of the residual noise 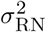 can be obtain as shown in Eq. S17.

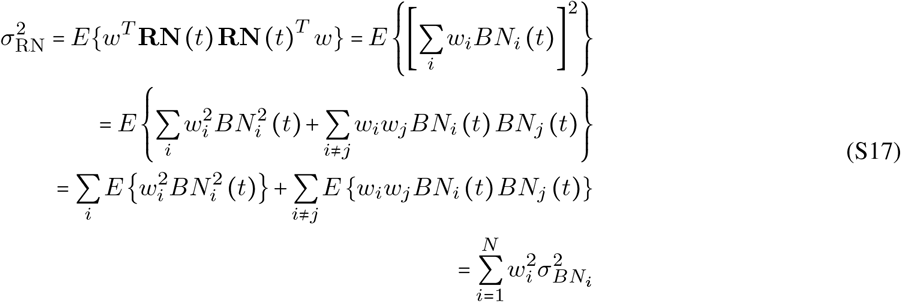

Now, if weights are defined as in Eq. S10, Eq. S17 can be written as

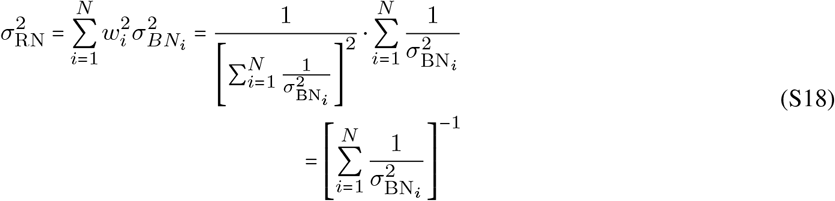

Note that Eq. S17 can be understood as the summation of *χ*2 distributed random variables with different weights (Eq. S5e), that is

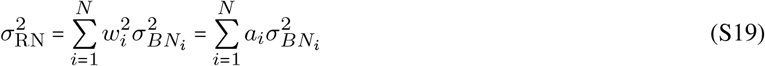

### Sup. 1.10. Estimation of effective number of epochs

Averaging the data using any kind of weights will affect the way in which each epoch contributes to the data, so that the effective number of epochs will be lower than the total number of epochs (or epochs) *N* . However, given that Eq. S19 can be interpreted as the linear combination of several variances with unknown DOFs, we can estimate the effective sampling size (or DOF) using the Welch-Satterthwaite approximation (Satterthwaite, 1941; Satterthwaite, 1946; Welch, 1947; Welch, 1956).

By means of the properties of a *χ*^2^ random variable (Eq. S5d), we can rewrite Eq. S19 and derive the DOF of a new distribution which shares the same first moment 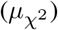 and the same variance 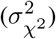, of 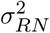.

First, let’s consider the case where 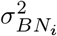 is estimated by averaging the variance *across epochs* (Eq. S15). If all samples are independent and drawn from the same random process, we can simplify and write the following

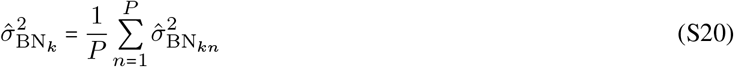

Now, using Eq. S5c we can rewrite the variance of each of the *n* sample points as

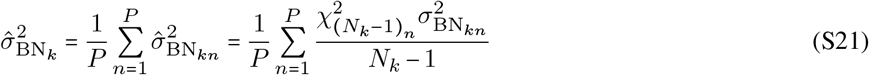

Keeping in mind that the noise is assumed to be a locally stationary ergodic processes (i.e., 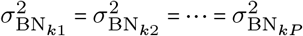) and 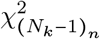 are *n* independent chi-squared variables with (*N*_*k*_ − 1) DOF we obtain

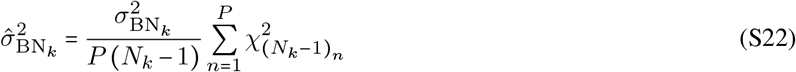

Next, considering the variance of the residual noise in the form of Eq. S19 combined with Eq. S22, we can rewrite 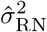 as

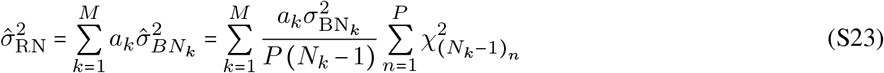

Using Eq. S5f, Eq. S23, and Eq. S5b, we can estimate the variance of our estimate—*var*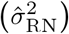—and represent it as the variance of a 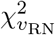 with unknown *v*_RN_ DOF

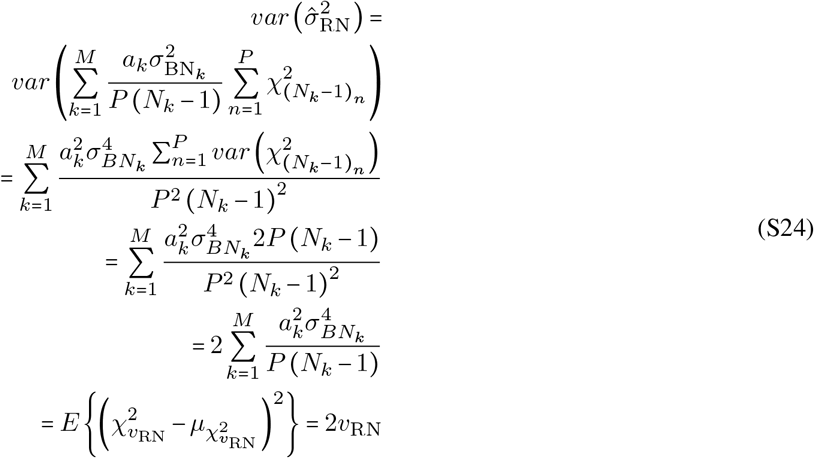

Similarly, recalling Eq. S5a we can estimate the mean (or expected value) of our estimate—*E* 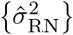—and express it as the expected value of a 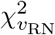 with unknown *v*_RN_ DOF, i.e.,

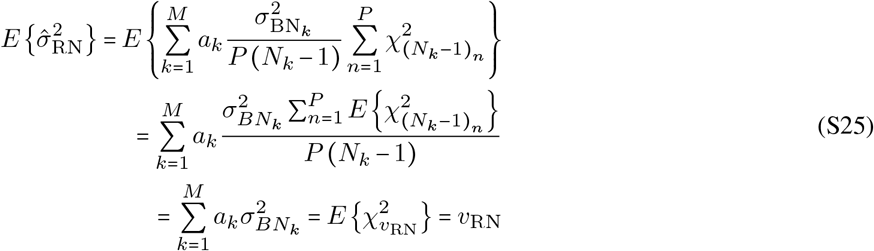

Squaring Eq. S25 and dividing by Eq. S24, the unknown DOF can be estimated as

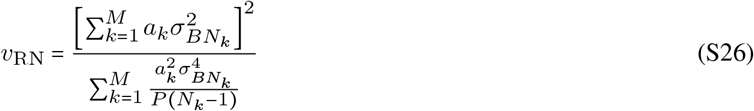

Replacing the constants *a*_*k*_ by 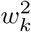, where *w*_*k*_ is given by Eq. S10, results in

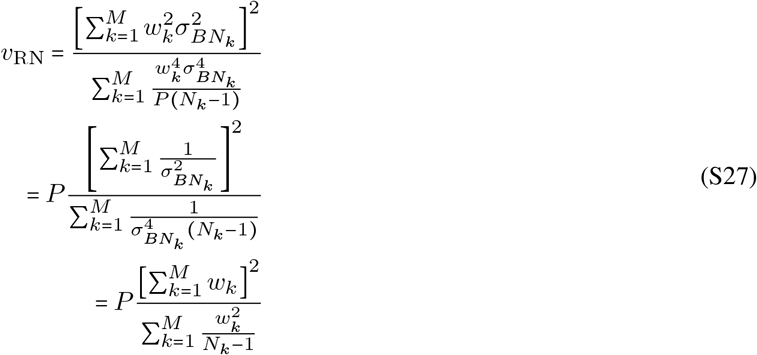

where *N*_*k*_ is the number of epochs used to estimate each variance 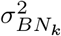, that is 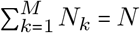, with M being the number blocks.

Finally, if the weights are expanded to all N individual epochs, and the number of epochs within a block is constant *N*_1_ = *N*_2_ = …. = *N*_*k*_, Eq. S27 can be approximated as

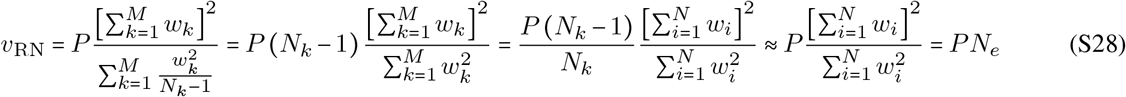

where *N*_*e*_ is the effective number of epochs estimated from the weights (*N*_*e*_ ≤ *N*).

Following the same rationale, when 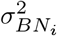 is estimated *within each epoch* according to Eq. S16, the DOF of 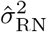 can be estimated as it follows

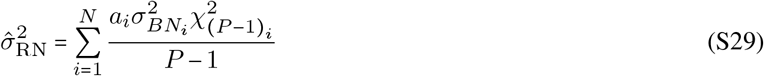

with variance

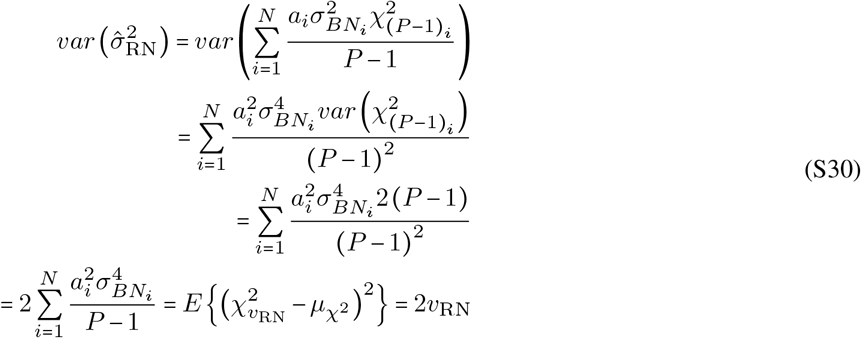

and expected value

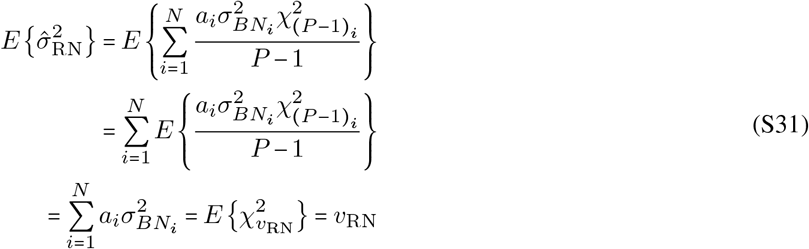

Squaring Eq. S31 and dividing by Eq. S30, followed by substituting *a*_*k*_ by 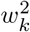 we obtain

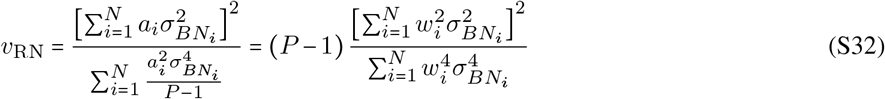

Finally, given that the weights correspond to Eq. S10, Eq. S32 can be rewritten as

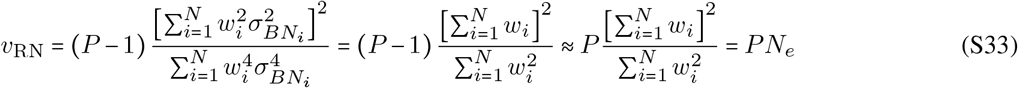

Equation S28 and Eq. S33 are essentially identical and provide an estimate of the *effective sample size* when weighted averaging is used. This approximation assumes that the samples used to estimate the variances are independent, within and across epochs. Independency across epochs can be approximated by removing low frequency components (i.e., high-pass filtering the raw data) with a cut-off frequency higher than 1/*T* . However, independency within epochs cannot be avoided either because of the properties of the background noise (coloured noise) and/or the use of non-ideal filters to reduce undesired noise. Therefore, the number of independent samples within epochs—*P* —used in Eq. S28 and Eq. S33, will be ≤ than the number of within-epoch samples used.

### Sup. 1.11. Estimation of the degrees of freedom within epoch

Consider a continuous time series in which the underlying noise is an ideal white noise, i.e., all frequency components have the same energy. In this ideal case, the number of DOF will be only limited by the bandwidth of the noise. In the case of a discretised white noise signal, the number of DOF will limited by the number of discretised samples for a full bandwidth noise, and it will be less than this if the bandwidth of the noise is lower than the Nyquist frequency of the recording system. However, the ongoing brain activity is typically dominated by low-frequency energy, reflected by an uneven distribution of its spectral power—coloured noise—usually described as being ∝ 1/*f*, with *f* being the frequency. A direct consequence of coloured noise is a higher level of correlation among samples which in turn reduces their independence as well as the effective DOF relative to a white noise.

This problem has been investigated in other fields and it forms the basis of the next steps to derive the effective DOF of the background neural activity present in each epoch (Donelan and Pierson, 1983; Medina et al., 1985; Young, 1986; Elgar, 1987; Rodríguez et al., 1999; Bendat and Piersol, 2010).

Consider a random variable *y*(*t*) with zero mean and standard deviation 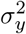. The autocorrelation function of this signal is given by

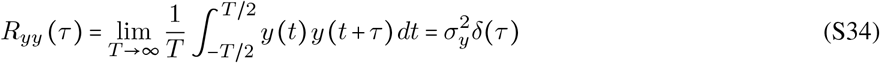

and its power spectral density is given by

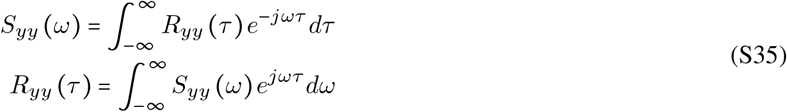

Note that

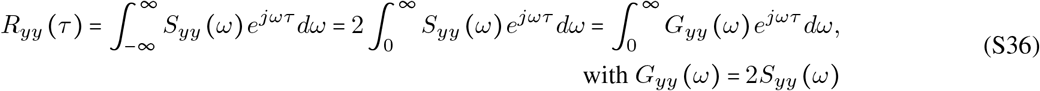

The power spectrum of an unlimited random process *y* (*t*) is given by

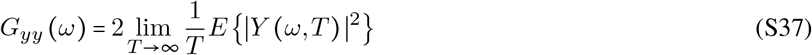

Therefore, an approximation for a time-limited signal is given by

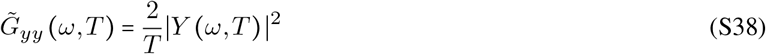

An important observation is that

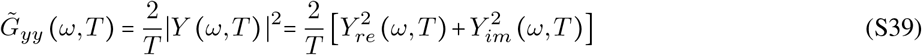

where the real and imaginary part are both normally distributed with

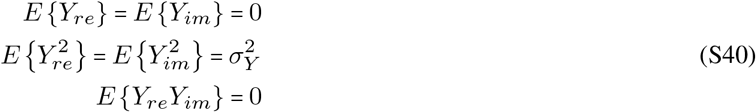

By definition, Eq. S39 follows a Rayleigh distribution, i.e., a *χ*^2^ distribution with two DOF, that is, 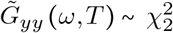. Therefore, using Eq. S5c, we can write

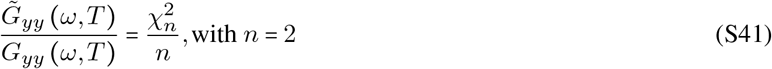

The normalised error of 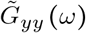 is given by

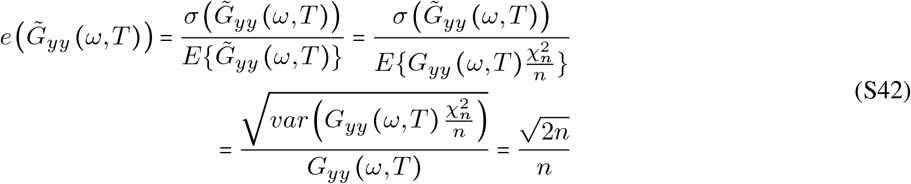

where the expected value and variance terms were derived using Eq. S5a and Eq. S5b, respectively.

Now, using Eq. S42, we can derive the variance of the estimated power spectrum as

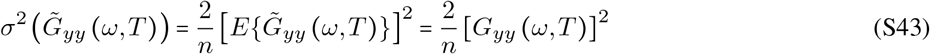

Thus, a better approximation of *G*_*yy*_ can be obtained by averaging several realisations of 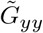

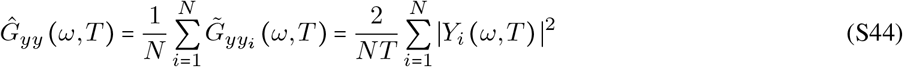

where the normalised error is now given by

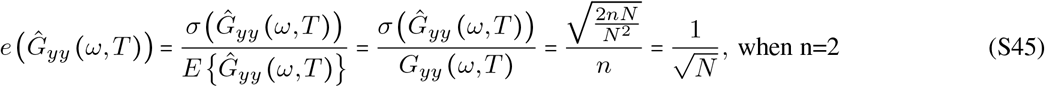

Note that Eq. S44 can be also estimated using weighted average via Eq. S8.

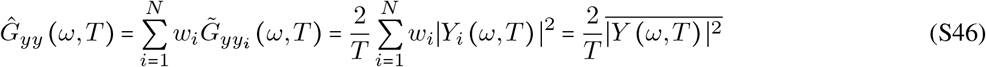

Although Eq. S46 represent a continuous function of *ω*, the following derivations will assume that we are working with a discrete representation, i.e., *ω*_*j*_, with *j* ∈ [1, *N*_*ω*_].

To derive the DOF of a signal, we can now invoke Parseval’s theorem which establishes that the power of a time-domain signal is equal to the sum of the power spectrum across frequencies, i.e., 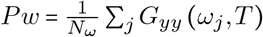, which for a zero-mean signal is equivalent to the variance of the signal.

Note that *G*_*yy*_ (*ω*_*j*_, *T*) follows a *χ*^2^ distribution, and so does its power ∑_*j*_ *G*_*yy*_ (*ω*_*j*_, *T*).

Consequently, can invoke once more Eq. S5d to derive the DOF for the signal power as

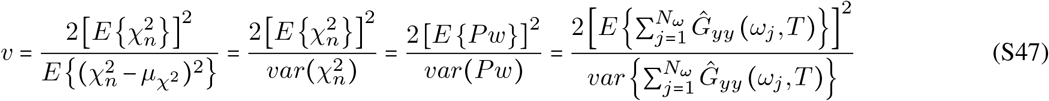

The expected mean of the power in Eq. S47 corresponds to

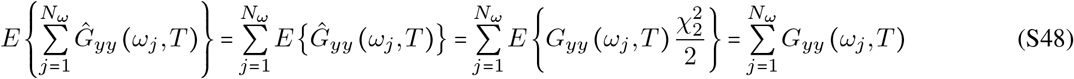

The variance in the denominator of Eq. S47 can be estimated using Eq. S43

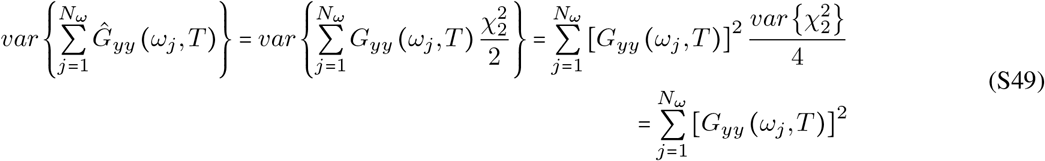

Replacing Eq. S49 in Eq. S47 we obtain

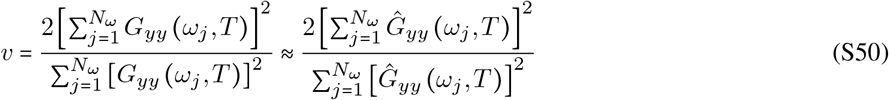

Eq. S50 thus provides an estimation for the DOF of the background activity within epochs of length *T*.

Note that the equivalent relationship in the continuum domain—useful to generate analytical solutions—is given by

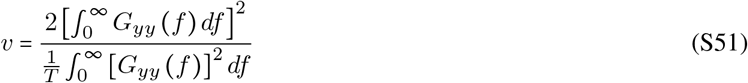

with *f* = *ω*/2*π* and *T* being the time interval analysed.

This equivalence becomes apparent when approximating the integral using the rectangle rule, i.e., replacing 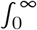 by 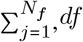 by Δ*f*, and *f* by *j*Δ*f*, with Δ*f =* 1/*T*.

In Eq. S51 can be seen that whilst *v* is shaped by the PSD function, it scales linearly as a function of *T*.

### Sup. 1.12. The Fmpi

As stated in Eq. S14, the statistical significance of the SNR is governed by the F-value, which in turn depends on the effective number of DOF *v*_*y*_ and *v*_RN_ of 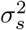 and 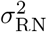, respectively. Different implementations based on the F-test (Elberling and Don, 1984; Elberling and Wahlgreen, 1985; Don and Elberling, 1994; Silva, 2009) have used empirical estimations of the degrees of freedom of 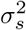—based on the background activity recorded from humans—, whilst it has been assumed that the DOF of 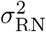 is given by the number of averaged epochs.

Whilst these approximations work for the average population, or an assumed worst-case scenario, they are unable to reflect the unique properties of the individual’s underlying noise that determines true value of those parameters.

The Fmpi algorithm exploits the information extracted from the underlying noise to accurately estimate the effective number of DOF of it (Eq. S50), tailored—or individualised—to the EEG acquired in the individual, and uses this information to estimate the effective number of DOF of 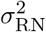. This is because Eq. S50 is an approximation of the effective number of independent samples within an epoch (*P* in Eq. S28 and Eq. S33).

### Sup. 1.12.1. Fmpi for time-domain analysis

Consider that we have epochs of length T, and we are interested in testing whether a signal is present within a shorter analysis window of duration *T* ^′^, with *T* ^′^ ≤ *T*.

To estimate the DOF of 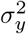—the variance within the analysis window—*v*_*y*_, we can make use of Eq. S50, which scales linearly as a function of the length of the time window chosen (Eq. S51).

Therefore, if 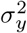 is computed from an analysis window *T* ^′^, the DOF (*v*_*y*_; the numerator of the F ratio) can be estimated from the entire window of duration *T* (i.e., at a higher frequency resolution), and then scaled down by Eq. S52

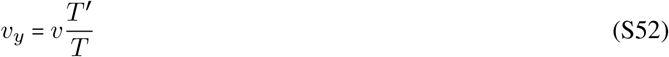

That is, *v* is estimated for the entire epoch of duration T using Eq. S50, with 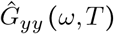 given by Eq. S46 (for discrete frequencies *ω*_*j*_), and where

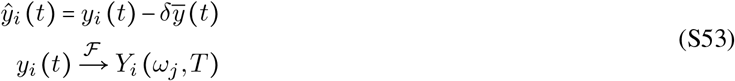

where ℱ denotes the Fourier transform and *δ* can be set to 0 if the power of the target response is much smaller than the background noise or 1 if the power of the target response is expected to be comparable to the background noise.

The DOF of 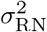 (*v*_RN_, the denominator of the F ratio) can be estimated by Eq. S54 for the entire epoch,

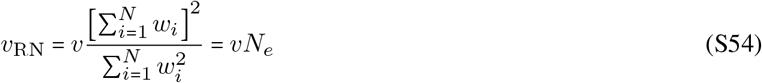

Thus, we can now compute the probability of F by

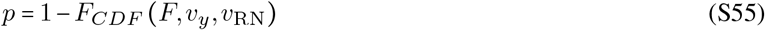

Where *F*_*CDF*_ (*F, v*_*y*_, *v*_RN_) is the cumulative distribution function of an F-distributed random variable with DOF *v*_*y*_ and *v*_RN_.

### Sup. 1.12.2. Fmpi in the frequency-domain—narrow-band frequency detection

There may be applications in which frequency-domain analysis is more appealing to detect the signal of interest. For example, if the stimulus evoking the response is presented periodically, such that the target response is assumed to also be a periodic signal, one may prefer to exploit this property and use a frequency-domain approach. The Fmpi, is well-suited for this purpose.

First, it is important noticing that the variance of a time-domain signal can be also estimated in the frequency domain by means of Parseval’s theorem. That is,

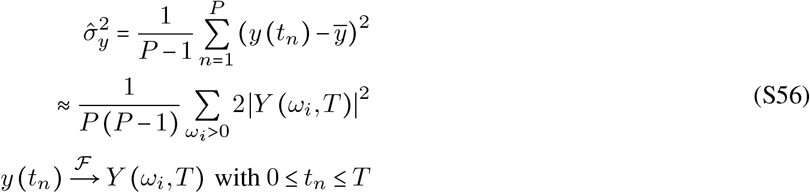

We can use this property to estimate the frequency-specific variance of the noise 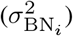 by summing the power spectral density components of the frequencies of interest as

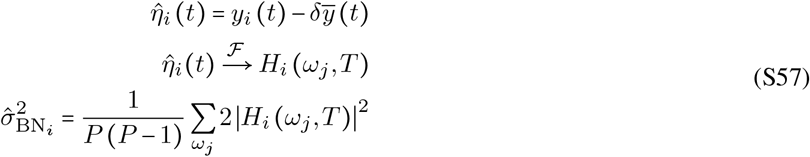

where *ω*_*j*_ ∈ [*w*_*a*_, *w*_*b*_, …, *w*_*m*_] is a set of frequencies of interest.

With that, we can estimate the frequency-specific residual noise using Eq. S17 and the F-ratio within a time window of duration *T* ^′^ (with P’ samples) as defined in Eq. S11, in which case the frequency-specific variance of the averaged response is given by

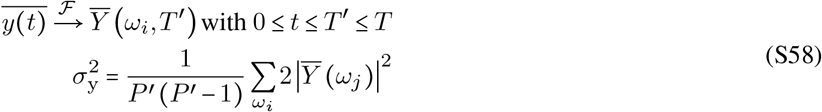

where again *ω*_*j*_ ∈ [*w*_*a*_, *w*_*b*_, …, *w*_*m*_].

The significance of the F-value depends on the DOF of 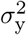 and 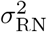.

Both *v*_*y*_ and *v*_RN_ can be estimated in a frequency-specific manner by Eq. S59 and Eq. S60

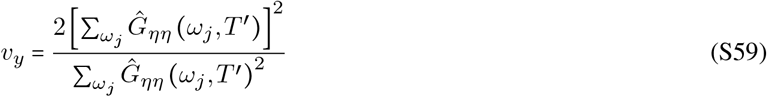

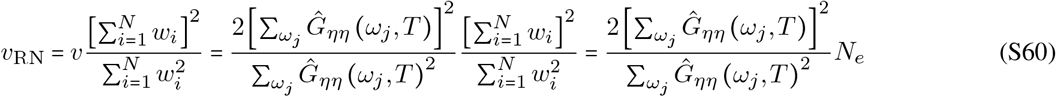

where *v* is similar to Eq. S59, but computed from the entire epoch of duration *T*, and the individuals weights *w*_*i*_ are the inverse of the frequency-specific variance (Eq. S57).

Note, however, that using different analysis windows for the estimation of the two variances (Eq. S57 and Eq. S58) will result in different frequency resolutions for each estimate. This can result in an imbalance between the energy contained within each frequency bin and the estimated F-value. This problem can be avoided employing the same analysis windows (*T = T* ^′^) for both estimates.

### Sup. 1.12.3. Fmpi in the frequency-domain—sparse frequency-detection

The above equations will work well if the number of samples used to estimate the background noise is large enough so that both the estimation of 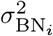 and *v*_RN_ are reliable. This is usually the case when analysing a narrow-band signal with a several frequency components, e.g., the range between 100 and 2000 Hz. However, for some applications we may be interested only in few frequency bins, e.g., the fundamental frequency and several harmonics, in which case the estimation of 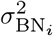 (using only a few frequency bins) will be poor.

To address this problem we can we can proceed as it follows. Consider that we have M sparsely distributed frequencies of interest *ω*_*j*_ ∈ [*w*_*a*_, *w*_*b*_, …, *w*_*m*_] . Next, assume that we want to assess if the pooled power of these frequency components is significantly larger than the underlying background noise for the same frequencies. Because of the low number of frequencies to be analysed—relative to the full spectrum of the data—the estimation of 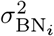 (Eq. S57) can be inaccurate. To avoid this problem, we can exploit the assumption that the noise at the frequencies of interest should be similar to that of their neighbouring frequencies. Under this assumption, we can obtain a good approximation of the power of the background noise at each frequency of interest by averaging the power of frequencies in their neighbourhood. That is,

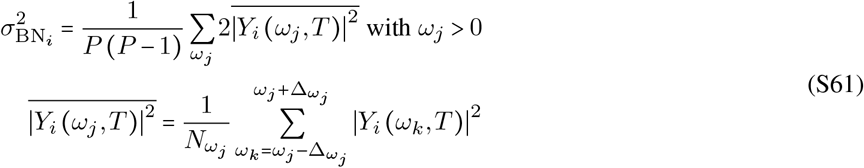

where 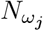 is the number of frequency bins around each frequency of interest used to estimate 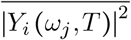. Note that in Eq. S61 the number of samples and, therefore, the number of DOF will be higher as more samples contributed to this estimation. The number of DOF given by Eq. S60 should now include all the frequencies used to estimate 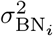, that is,

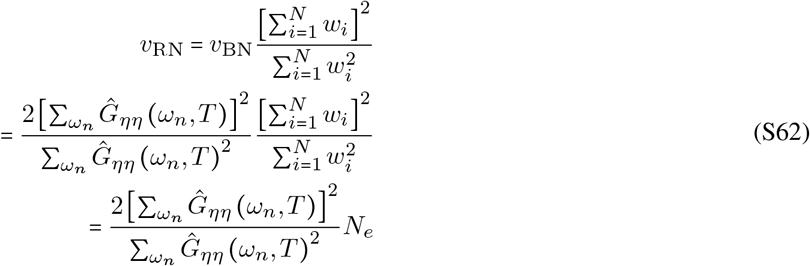

with *ω*_*n*_ ∈ [*ω*_1_ −Δ*ω*_1_, …, *ω*_1_ + Δ*ω*_1_, *ω*_2_ −Δ*ω*_2_, …, *ω*_2_ + Δ*ω*_2_, …, *ω*_*m*_ − Δ*ω*_*m*_, …, *ω*_*m*_ + Δ*ω*_*m*_].

Finally, we can estimate 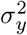 and *v*_*y*_ by means of equations Eq. S58 and Eq. S59 and the probability of the F-value by Eq. S55.

### Sup. 1.12.4. Predicting the necessary number of epochs to detect a response

An estimation of the necessary number of epochs to detect a response can be derived by taking into account the relation between Eq. S6 and the RN as a function of the number of epochs.

An EP will be detected whenever the ongoing *F value* becomes greater than the critical *F* − *value* (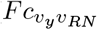, Eq. S63), that is, the threshold to reject the null-hypothesis.

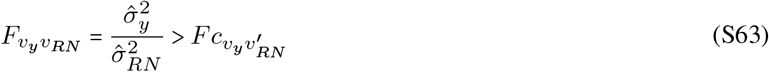

Here, the subscript *v*_*y*_ indicate the DOF obtained by averaging *N* epochs and 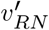 indicates the expected DOF at the target number of epochs. Next, assuming independency across epochs, we can estimate variance of the RN as a function of the number of epochs 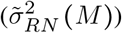 and the ‘equivalent’ background noise variance 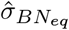 by Eq. S64

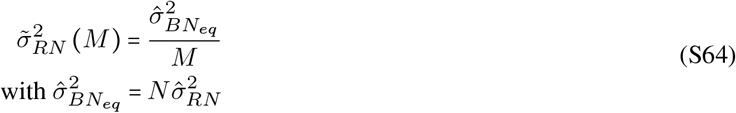

where *M* is the arbitrary number of epochs.

Using Eq. S64 in Eq. S63, we can estimate the total number of necessary epochs to detect an EP, *N*_*s*_, by

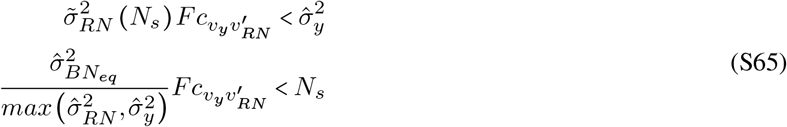

The denominator on the left side of Eq. S65 reflects the fact that the variance of the average response consists of the variance of the neural response 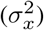 and the variance of the RN, i.e., 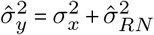, and in the absence of an EP it should not be lower than the variance of the RN. That is, *N*_*s*_ cannot exceed the upper limit given by

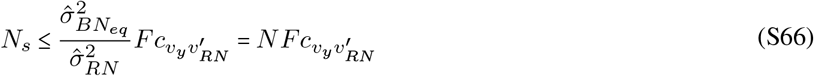

The DOF *v*_*y*_ is given by Eq. S52 or Eq. S59, and 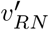 is derived from the current *v*_*RN*_ projected at *N*_*s*_ as

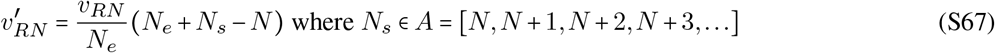

Here, *N*_*s*_ is *min*(*A*) for which Eq. S65 satisfied.

### Sup. 1.13. Analytical DOF estimation

Consider a noise with a PSD that follows a 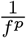 function, with *f* > 0 being the frequency. The PSD of the noise passed through a linear filter—*Y*_*yy*_—is given by

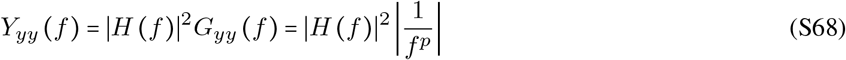

Now, consider an ideal bandpass filter (Eq. S69) and the continuous expression of the DOF given by Eq. S51

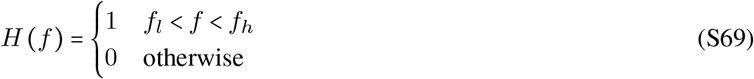

We can obtain the analytical DOF for bandpass-filtered 1/*f*^*p*^ PSD noise within a given analysis time window of duration *T* as

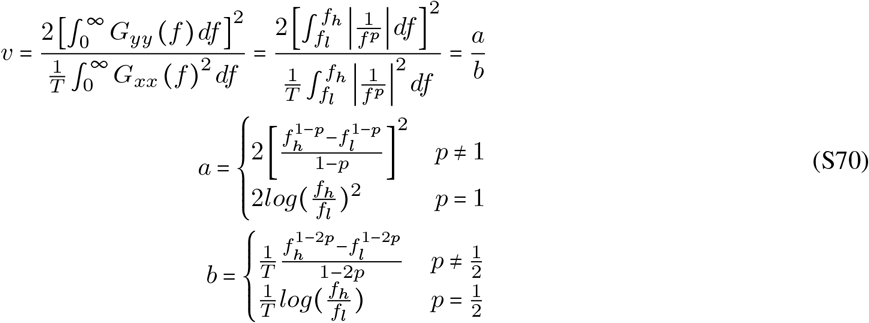

## Supplementary figures

**Sup. Figure 1.**
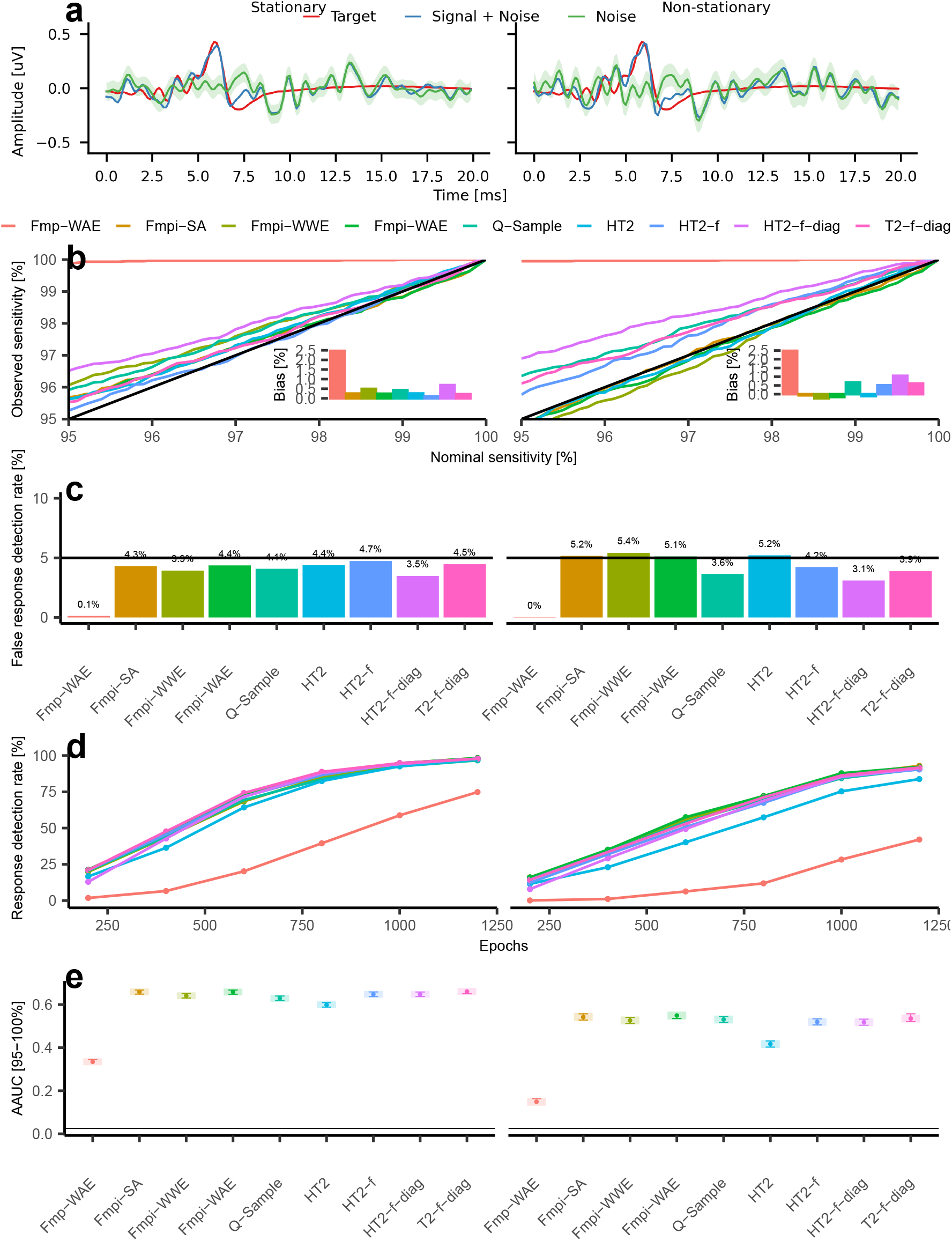
Fmpi enables robust performance under stationary and non-stationary noise in brainstem response simulations. **a** Examples of ABR simulations under stationary (left) and non-stationary (right) background noise. The template ABR (Target) is shown in red. Weighted average waveforms containing both the ABR template and noise are depicted in light blue (Signal + Noise), whilst averages obtained without the target ABR (Noise) are shown in green, with the standard error of the mean indicated by the shaded green area. **b** Nominal versus observed sensitivity for clinically relevant thresholds (95-100%), generated by pooling noise-only data across all epochs. The diagonal solid black line represents the ideal unbiased expectation. Insets show the average bias across nominal sensitivities; a positive bias indicates higher observed sensitivity than nominal, whilst a negative bias indicates the opposite. **c** False response detection rates from the noise-only data at a nominal sensitivity of 95%, pooled across all epochs. **d** Response detection rates (Signal + Noise) as a function of the number of epochs. Each epoch includes 1000 simulations. **e** AAUC for the clinically relevant nominal sensitivities. Shaded areas represent 95% confidence intervals. Error bar lengths have been adjusted for visual post hoc comparisons (Holm FWER corrected); non-overlapping segments indicate significant differences, whilst overlapping segments indicate non-significant differences. The horizontal solid black line indicates the chance level of the AAUC. The different methods are indicated by colour across all panels.

**Sup. Figure 2.**
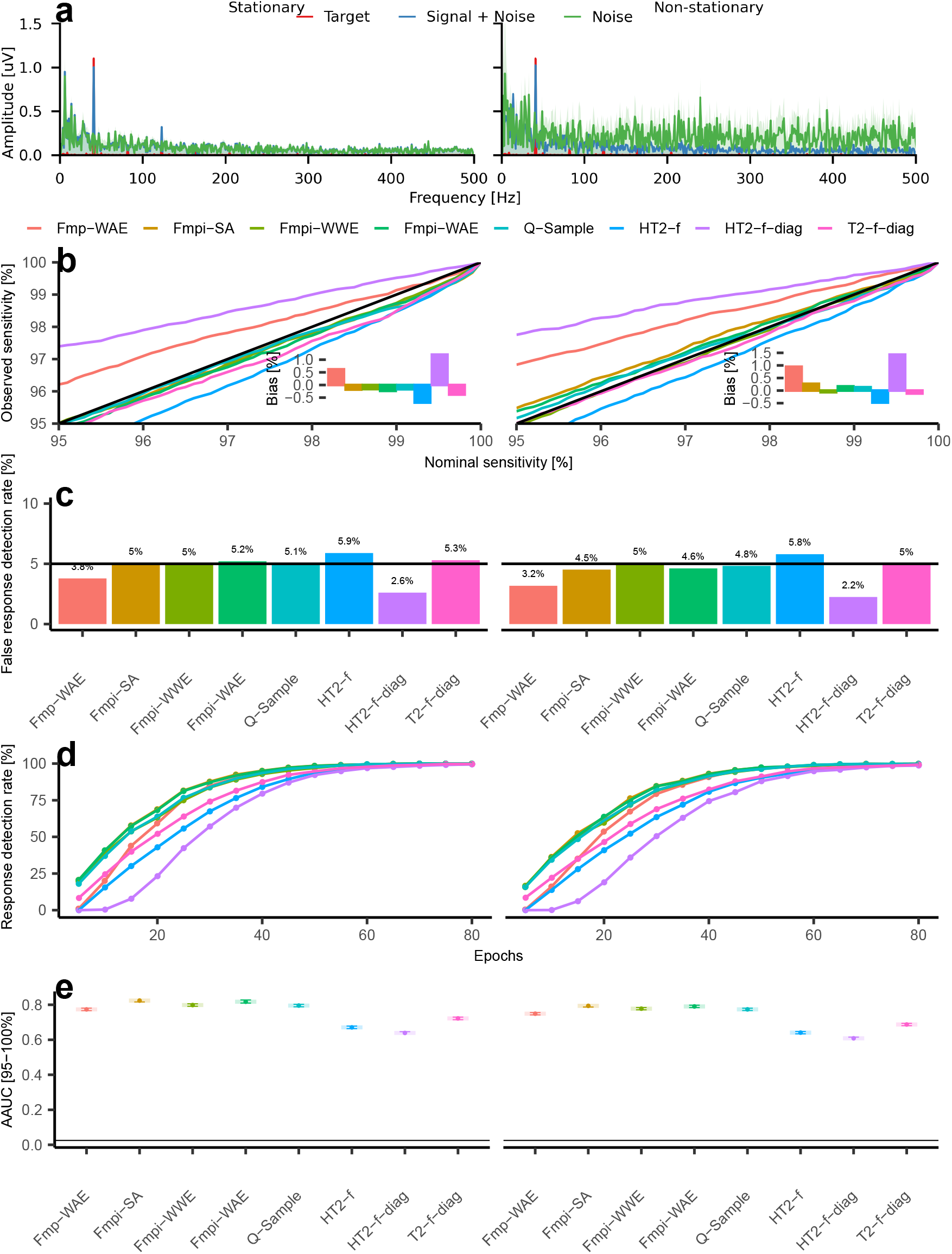
Fmpi enables robust performance under stationary and non-stationary noise in steady-state response simulations. **a** Examples of 41 Hz ASSR simulations under stationary (left) and non-stationary (right) background noise. The template ASSR (Target) is shown in red. Weighted average waveforms containing both the ASSR template and noise are depicted in light blue (Signal + Noise), whilst averages obtained without the target ASSR (Noise) are shown in green, with the standard error of the mean indicated by the shaded green area. All detectors employed 3 harmonics (six features) for statistical assessments. **b** Nominal versus observed sensitivity for clinically relevant thresholds (95-100%), generated by pooling noise-only data across all epochs. The diagonal solid black line represents the ideal unbiased expectation. Insets show the average bias across nominal sensitivities; a positive bias indicates higher observed sensitivity than nominal, whilst a negative bias indicates the opposite. **c** False response detection rates from the noise-only data at a nominal sensitivity of 95%, pooled across all epochs. **d** Response detection rates (Signal + Noise) as a function of the number of epochs. Each epoch includes 1000 simulations. **e** AAUC for the clinically relevant nominal sensitivities. Shaded areas represent 95% confidence intervals. Error bar lengths have been adjusted for visual post hoc comparisons (Holm FWER corrected); non-overlapping segments indicate significant differences, whilst overlapping segments indicate non-significant differences. The horizontal solid black line indicates the chance level of the AAUC. The different methods are indicated by colour across all panels.

**Sup. Figure 3.**
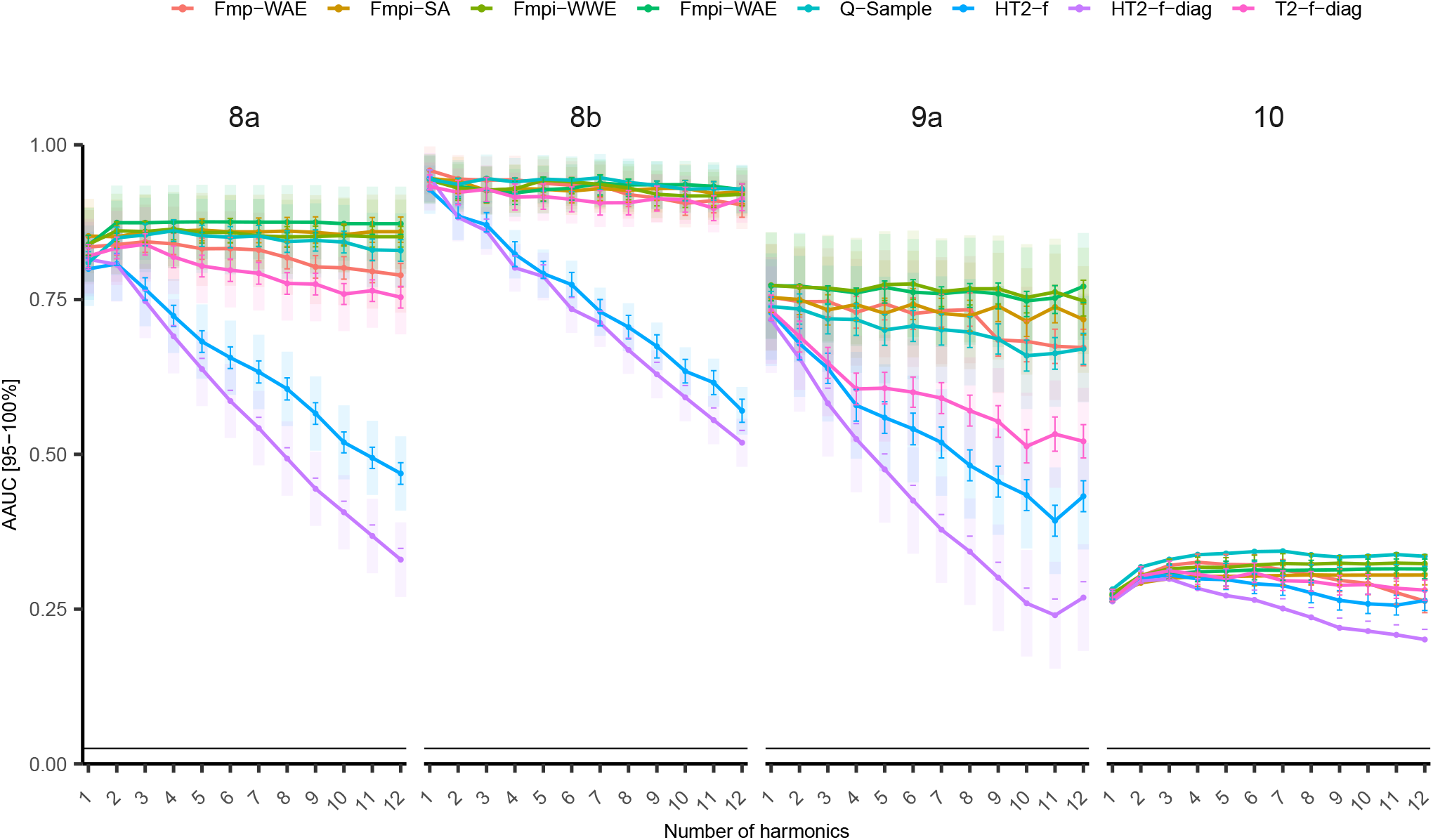
Fmpi enables robust performance for human steady-state response analyses in the frequency domain. ASSRs from different studies (each column represents a different study) obtained for ASSR rates of ≈ 40 and 81 Hz (study 8), 81 Hz (study 9), and 38, 68 and 69 Hz, simultaneously (study 10). The AAUC for the clinically relevant nominal sensitivities is shown as a function of the number of harmonics included in the analyses, spanning from 1 to 12 harmonics. Shaded areas correspond to 95% confidence intervals. The length of the error bars has been adjusted for visual post hoc comparisons (Holm FWER corrected). Non-overlapping segments indicate significant differences, whilst overlapping segments indicate non-significantly differences. The horizontal solid black line indicates the chance level of the AAUC. The different methods are indicated by the colours across all panels.

**Sup. Figure 4.**
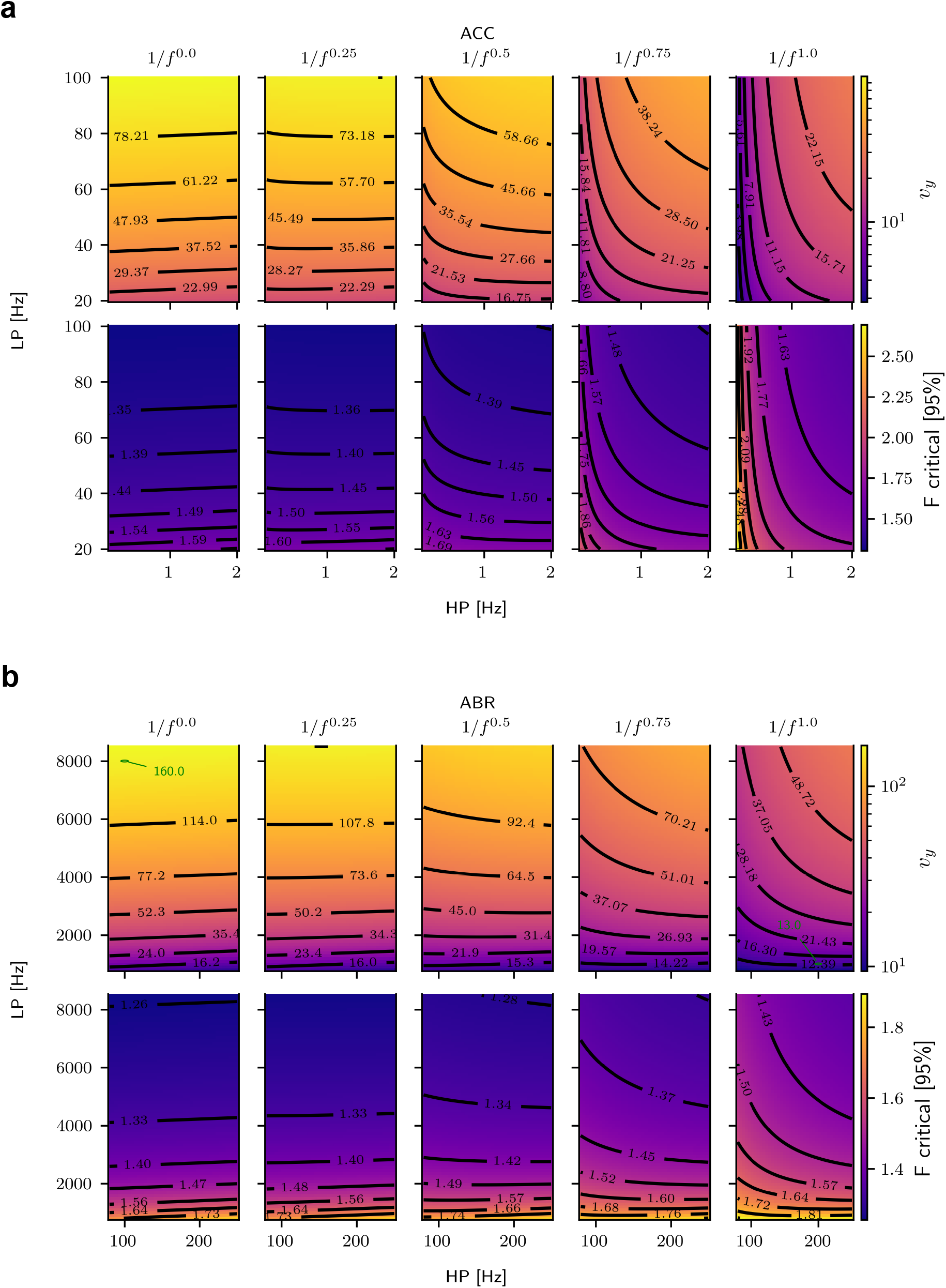
Analytical DOF and critical F-values for various stationary noise types and bandwidths. **a** DOF *v*_*y*_ and 95% critical F-value for a fixed *v*_*RN*_ = 256 for cortical-like analysis bandwidths, using an analysis window of 500 ms. The low-pass (LP) and high-pass (HP) ideal cutoff frequencies are indicated on the y- and x-axes, respectively. Each panel corresponds to a noise with a PSD of 1/*f*^*p*^ with *p* ranging from 0 (white noise) to 1 (pink noise). The range of the corresponding values is shown by the colour bar. **b** Same as in panel **a**, but for ABR-like analysis bandwidths, using an analysis window of 10 ms. The green markers for white (p=0) and pink (p=1) noise in panel **b** correspond to analytical DOF values of *v*_*y*_ = 160.0 (white noise) and *v*_*y*_ = 13.0 (pink noise), respectively. The empirical values reported by Elberling and Don (1984) for the same bandwidth and noise types were 160 and 15, respectively.

